# Predicting COVID-19 hospitalisation and common disease risk from comorbid diagnoses in 13 million individuals

**DOI:** 10.64898/2026.08.27.26361302

**Authors:** Hongjiao Liu, Mehrdad A. Mizani, Yujie Zhao, Angela Wood, Michael Inouye, Alkes L. Price, Xilin Jiang, the CVD-COVID-UK/COVID-IMPACT Consortium

## Abstract

Predicting disease risk from prior diagnoses is fundamental to clinical decision-making, particularly during health emergencies such as the COVID-19 pandemic, when individuals with long-term conditions may be disproportionately vulnerable to adverse outcomes. Despite intense interest in developing models to predict disease risk from prior diagnoses ^1–3^, most prediction models do not estimate effects of each prior diagnosis on disease risk conditional on other diagnoses, limiting interpretability and clinical utility. We developed the Comorbidity Risk Score (CRS), trained on 13 million individuals (age 40-69) from linked electronic health record (EHR) datasets of the entire population of England, to predict COVID-19 hospitalisation and 87 other disease outcomes. CRS was trained at close to saturated sample size and precisely estimated the effects of 212 prior diagnoses on the 88 disease outcomes, conditional on all other prior diagnoses. Correlations of CRS effect sizes across outcomes (e.g. 0.76 for myocardial infarction vs. hyperlipidaemia) matched the corresponding genetic correlations (e.g. 0.79 for myocardial infarction vs. hyperlipidaemia), confirming that comorbidity architectures capture disease aetiology. On average, CRS identified 5% of the population with 3.4-fold higher disease risk, including myocardial infarction (4.4-fold), lung cancer (6.5-fold), and COVID-19 hospitalisation (6.3-fold). Using prior diagnoses alone, CRS outperformed state-of-the-art clinical COVID-19 models ^4^. Furthermore, CRS (*N*=13 million) substantially outperformed state-of-the-art AI ^1^ (*N*=0.5 million) and linear ^3^ (*N*=0.5 million) models in predicting disease risk, suggesting that training sample size outweighs model complexity. CRS attained near-perfect transferability across self-reported ethnicities (e.g., Black vs. White: AUROC ratio = 97.3%). Finally, CRS distinguished independently predictive comorbidities from indirect associations, e.g., lipid metabolism disorder was a strong predictor of myocardial infarction risk but not ischaemic stroke, after conditioning on other prior diagnoses. In conclusion, CRS provides a comprehensive resource for understanding the impact of comorbidities on COVID-19 and other future diseases, revealing disease aetiology while enabling powerful prediction of disease risk.

## Introduction

Prior diagnoses carry important information about future disease risk and are crucial for clinical decision making. Understanding the risks associated with prior diagnoses becomes especially useful during health emergencies such as the COVID-19 pandemic, when individuals with long-term conditions may face disproportionate risk of adverse outcomes. In particular, the impact of comorbidities on severe COVID-19 has been long recognised ^5–7^. Recent studies have developed models using longitudinal electronic health records (EHRs) for diagnoses-based disease risk prediction ^1–3^, facilitating clinical risk stratification and targeted prevention with the potential of improving long-term patient outcomes. However, these models do not estimate the effect of each prior diagnosis on future diseases conditional on other diagnoses, which limits interpretability and clinical utility. Precise estimates of prior-diagnosis effects on future diseases are directly informative for clinical management of diseases with high public health burden, including cardiovascular disease ^8^, type 2 diabetes (T2D) ^9^, and severe COVID-19 (e.g., the QCovid model) ^4,10^.

Many efforts have been focused on developing machine learning or artificial intelligence (AI) models for EHR data. Earlier studies derived EHR-based machine learning models to predict individual diseases or outcomes such as pancreatic cancer ^2^, coronary artery disease ^11^, and mortality in intensive care units ^12^. More recently, generative AI models have been adopted to simultaneously predict a wide range of disease outcomes ^1,13,14^. For example, utilising generative pre-trained transformers (GPTs), Delphi-2M predicts over 1,000 diseases based on past disease history from UK Biobank EHR data ^1^. On the other hand, in a similar vein to polygenic risk scores (PRSs), Detrois et al. developed phenotype risk scores (PheRS) for 13 diseases via elastic net models that are transferable across biobanks ^3^. Despite the ability of AI models to capture complex relationships within EHR data, these models are usually trained with limited sample sizes and do not estimate the effect of each prior diagnosis, hence limiting interpretability and utility in routine clinical settings.

In this study, we developed the Comorbidity Risk Scores (CRSs), a generalised linear risk model trained and validated on 13 million individuals from the CVD-COVID-UK cohort, an EHR database covering the entire population of England, to predict COVID-19 hospitalisation and 87 other common disease outcomes. Leveraging a nation-wide sample size, we precisely estimated the conditional effect sizes of 212 prior diagnoses on the disease outcomes, providing a comprehensive resource for the impact of comorbidities on future diseases. We demonstrated the strength of very large training samples by showing that CRS was highly predictive of future diseases, outperformed state-of-the-art AI models, and transferred well across ethnicity groups.

### Overview of comorbidity risk scores

We constructed comorbidity risk scores (CRSs) to predict future disease from prior comorbidities (Fig. S1a), using 13 million individuals aged 40-69 from the CVD-COVID-UK cohort ^15^ (average follow-up since first diagnosis = 23.5 years). We focused on this age group for its high risk of COVID-19-associated morbidity and mortality as well as its higher comorbidity burden ^16,17^. CRS predicts 5-year risk for COVID-19 hospitalisation and 10-year risk for all-cause mortality and 86 other common diseases (average number of training cases = 207,806; Table S1). CRS incorporates 212 predictor diseases (prevalence ≥ 0.2%), which we derived by aggregating 391 Phecodes into parent-level Phecodes to reduce collinearity (Table S2); these predictors accounted for 85.8% of all disease diagnoses in the EHR data. For each outcome and predictor disease, we used the earliest recorded diagnosis per individual from primary care and inpatient hospital data. CRS used only predictor diseases diagnosed any time three months before the start of outcome follow-up (i.e., index dates), which were 1 January 2020 for COVID-19 hospitalisation and 1 January 2015 for the other 87 outcomes (Methods).

CRS estimates the effect of each predictor on future disease outcomes conditional on all other predictor diseases. It is designed to capture the long-term effects of comorbidities across the full disease spectrum, rather than associations between diseases within the same category that tend to be diagnosed together. First, we excluded prior diseases from the same Phecode category as the outcome (Methods) to avoid including predictor diseases too closely related to the target outcome. For example, prior ischaemic heart disease (Phecode 411), such as angina pectoris and coronary atherosclerosis, was excluded from the CRS predicting myocardial infarction (Phecode 411.2). Second, we included a 3-month gap between predictor diagnoses and outcome index dates to avoid predicting incident events recorded soon after prior diagnoses (also see below for other time gaps we tested). Finally, we used inverse probability of censoring weighting (IPCW) ^18^ to account for individuals who were lost to follow-up (i.e., died) before the end of the 5/10-year period (Methods).

CRS was first trained in self-reported White British individuals born in odd years (*N* = 6,527,585) and tested in self-reported White British individuals born in even years (*N* = 6,511,145), as well as in all individuals, regardless of birth year, who self-reported as Asian (*N* = 961,625), Black (*N* = 502,070), Mixed (*N* = 145,980) and Irish/Other White (*N* = 998,920; Fig. S1c). As our studying cohort overlaps with other cohorts that contain participants from England (e.g., UK Biobank ^19^), this design allows users to validate CRS in those cohorts, where held-out samples can be defined by birth year. We also released CRS effect size estimates trained in all 13 million White British individuals (*N* = 13,038,730; Table S3).

We benchmarked CRS against state-of-the-art AI models trained in 0.5 million samples (Delphi-2M; tested outside CVD-COVID-UK) ^1^, PheRS ^3^, QCovid ^10^, and clinical comorbidity indices ^20^, as well as socioeconomic status and an age-and-sex baseline. We considered six performance metrics: fold change in cumulative risk for the top 5% of predictions (vs. population average risk), area under the receiver operating characteristic curve (AUROC), liability-scale *R*^2^, observed-scale *R*^2^, C-index, and area under the precision-recall curve (AUPRC) (Methods). We primarily focus on the first three metrics: fold change in risk at top 5% CRS distribution, AUROC, and liability-scale *R*^2^; however, in comparisons to other methods, we focus on ΔAUROC (vs. an age-and-sex baseline model) for consistency with the primary metric reported by Delphi-2M and PheRS.

### CRSs achieve powerful prediction for COVID-19 hospitalisation and related outcomes

CRSs were significantly more predictive than the age-and-sex baseline for 86 out of 88 outcomes (*P* < 0.001) across six metrics (Fig. 1a, Fig. S2, Table S4). Averaged across outcomes, CRS identified 3.4-fold higher disease risk at top 5% of the score distribution (1.8-fold improvement vs. Age + Sex), with an AUROC of 0.678 (ΔAUROC = 0.060 ± 0.000 vs. Age + Sex), C-index of 0.666 (ΔC-index = 0.057 ± 0.000), liability-scale *R*^2^ of 0.090 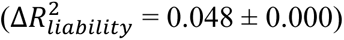, observed-scale *R*^2^ of 0.074 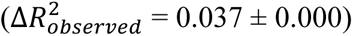, and AUPRC of 0.078 (ΔAUPRC = 0.023 ± 0.000). Throughout the manuscript, we focus on COVID-19 hospitalisation and four COVID-19-related disease outcomes (death, myocardial infarction ^21^, T2D ^22^, and lung cancer ^23^), while presenting results for all 88 disease outcomes in the Supplementary Tables. CRSs were strongly predictive (AUROC > 0.700) for 27 outcomes, including COVID-19 hospitalisation, death, and lung cancer (AUROC = 0.732-0.798), while COVID-19 hospitalisation, death, and T2D also showed large improvement over the age-and-sex baseline (ΔAUROC = 0.097-0.153). Individuals in the top 5% of the CRS distribution for COVID-19 hospitalisation, death, and myocardial infarction had 6.3-, 5.5- and 4.4-fold increased risk, respectively, compared to population-level risk. The predictive power of CRS was retained among individuals with the same number of prior diseases and increased with the number of prior diseases (Fig. S3), highlighting its value for managing multiple long-term conditions.

**Fig. 1:**
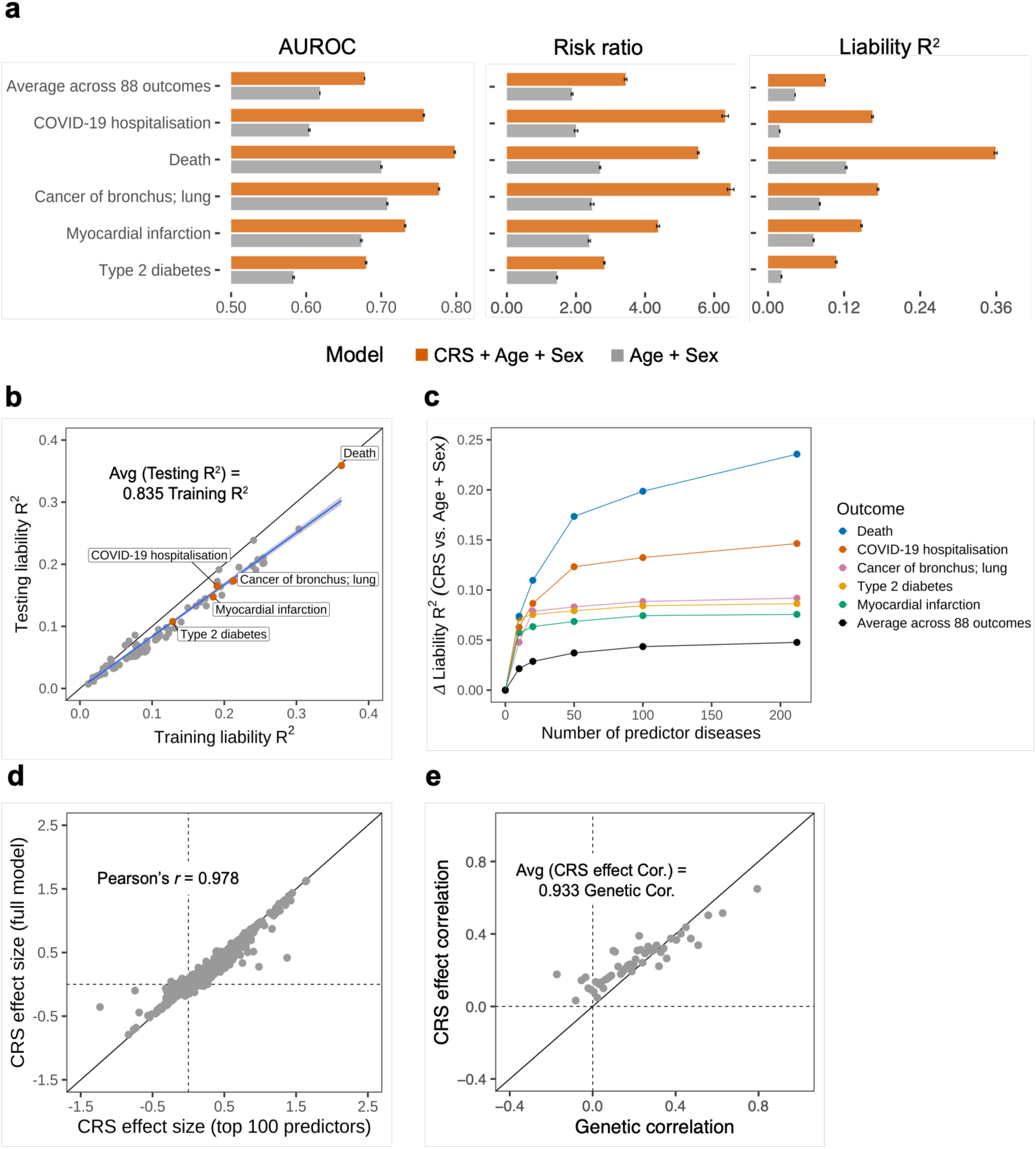
CRSs are highly predictive of common diseases. **a.** AUROC, observed risk ratio (top 5% of the predicted risk distribution vs. population average), and liability-scale *R*^2^ for example outcomes for the CRS model (including age and sex as covariates), compared to a baseline age-and-sex model. Error bars represent 95% CI around the performance metrics (but are generally very narrow). **b.** Liability *R*^2^ in the testing data (*y*-axis) vs. liability *R*^2^ in the training data (*x*-axis) for 88 outcomes, with example outcomes labelled and highlighted in red points. A linear regression through the origin is fitted (blue line) with 95% confidence bands, with the slope reported in text. **c**. Improvement in liability *R*^2^ by CRS over the age-and-sex baseline (Δ liability *R*^2^; *y*-axis) vs. number of predictor diseases included in the CRS model (*x*-axis), grouped by example outcomes. **d.** Effect sizes (log odds ratio) of the full CRS model with 212 predictors (*y*-axis) vs. effect sizes of the CRS model with top 100 prevalent predictors (*x*-axis). Each point represents the effect size for a unique predictor-outcome pair, with Pearson’s correlation between *x*- and *y*-axis reported in text. **e.** Correlation in CRS effect sizes across outcomes vs. genetic correlation across outcomes. For better visualisation, the pairwise genetic correlation was divided into 50 equal quantiles, and the mean genetic correlation within each quantile (*x*-axis) is plotted against the mean CRS effect correlation within each quantile (*y*-axis). The slope from linear regression through the origin of CRS effect correlation on genetic correlation across all pairs of outcomes (instead of quantile means) is reported in text. Source data are reported in Table S4 (Panel a), Table S10 (Panel b), Table S11 (Panel c), Table S12 (Panel d), and Table S6 (Panel e).

CRSs were trained at nearly saturated sample sizes. As training sample sizes increase towards infinity, regression models are expected to overfit less, resulting in converging training *R*^2^ and testing *R*^2^. At a training sample size of 6.5 million, the testing *R*^2^ of CRSs was 83.5% of the training *R*^2^ (Fig. 1b), indicating that most linear variance was captured at this sample size. We note that CRSs trained on the full 13 million individuals are expected to capture even more variance (Table S3). The large training sample also enables the direct estimation of comorbidity effects on future disease outcomes conditional on all other predictor diseases, without the need for regularisation (e.g. LASSO ^24^ or ridge regression ^25^). Consistent with this, we applied elastic net regression ^26^ to example diseases and determined that CRS achieved almost identical prediction accuracy to the elastic net model (Fig. S4). In total, CRS estimated 18,497 comorbidity effects (all with small standard errors (SE); average SE = 0.038 for CRS trained on 6.5 million samples, average SE = 0.027 for CRS trained on 13 million samples), of which 8,920 (48.2%) were significantly different from zero (FDR-adjusted P-value < 0.05, Table S5).

CRSs included a sufficient number of predictor diseases to capture most prior comorbidity information. To assess whether increasing the number of predictor diseases would improve CRS performance, we separately trained CRS models using different numbers of the most prevalent predictors (*N*_pred_ = 10, 20, 50, 100, 212; Fig. 1c). CRSs trained with the 100 most prevalent predictor diseases achieved similar accuracy (average *R*^2^ = 0.086) to the full model trained with 212 predictor diseases (average *R*^2^ = 0.090), suggesting that further increasing the number of predictor diseases is unlikely to improve performance. CRS effect sizes remained stable when increasing the number of predictor diseases from 100 to 212 (Fig. 1d), suggesting that sufficient covariate disease information has been conditioned on. Increasing the number of predictor diseases further from 212 to 400 would mostly add rare comorbidities (average prevalence = 0.08%), which reduces the precision of estimating conditional predictor effects (Fig. S5). Taken together, these results indicate that 212 predictor diseases are sufficient to capture most comorbidity information while providing adequate power to distinguish independently predictive prior comorbidities from indirect associations.

### CRS effect correlations match genetic correlations

Diseases are diagnosed with error; therefore, phenotypic correlations between disease outcomes cannot fully capture their shared aetiology. However, the correlation in CRS effect sizes between two disease outcomes (CRS effect correlation) captures their shared comorbidity profile and, given a sufficient training sample size, is not affected by diagnostic error (Methods). Out of 3,802 eligible outcome pairs (excluding parent and child Phecode pairs), 115 outcome pairs showed high correlations of CRS effect sizes (Pearson’s *r* > 0.7, Table S6), whereas only two pairs of disease outcomes had a phenotypic correlation above 0.5. On the other hand, genetic correlation quantifies the correlation of causal genetic effect sizes between trait pairs, reflecting the extent to which diseases share biological pathways at the molecular level ^27^. To assess how well CRS effect correlation captures shared disease aetiology, we computed genetic correlations among 44 of the 88 disease outcomes that have heritability *z*-score above 6 (Methods) and found that CRS effect correlations were highly consistent with genetic correlations, with a slope of 0.93 and a noise-adjusted correlation of 0.53 (Fig. 1e). On the contrary, the slope of phenotypic correlation on genetic correlation was 0.09 (Fig. S6). For example, myocardial infarction and hyperlipidaemia had a CRS effect correlation of 0.76 ± 0.03 and a genetic correlation of 0.79 ± 0.06, whereas their phenotypic correlation was only 0.13.

CRS effect sizes are particularly useful for studying shared disease mechanisms among diseases that are largely determined by environmental factors and therefore have limited genetic signals. For example, pneumonia, COVID-19 hospitalisation, and sepsis have limited reported heritability but are clinically correlated diseases ^28–30^, consistent with the high average CRS correlation among these outcomes (0.77), implying strongly shared disease mechanisms. In contrast, the average phenotypic correlation among these outcomes was only 0.21 (Table S6), suggesting that these shared mechanisms cannot be identified by simply computing phenotypic correlations. Mapping observed-scale phenotypic correlations to the liability scale increased the correlation strength but still captured less disease similarity than CRS effect correlations (Methods; Fig. S6, Table S6).

### CRSs outperform state-of-the-art models in predicting COVID-19 hospitalisation and related outcomes

CRSs aggregate the linear effects of prior comorbidities to predict future disease outcomes, whereas AI models capture nonlinear relationships among comorbidities for the same task ^1,2^. We compared the predictive accuracy of CRS (using prior diagnoses outside the target disease category, *N* = 13 million) with that of Delphi-2M ^1^, a transformer model trained in UK Biobank using all prior diagnoses, BMI, and smoking (*N* = 0.5 million) (Fig. 2a). Across the 49 overlapping outcomes, the average ΔAUROC for CRS (0.082 ± 0.001) was 33% higher than that of Delphi-2M (0.061). CRS outperformed Delphi-2M for 39 of the 49 outcomes, including myocardial infarction (ΔAUROC = 0.090 for CRS vs. 0.013 for Delphi-2M) and death (ΔAUROC = 0.112 for CRS vs. 0.097 for Delphi-2M). Delphi-2M performed better for obesity (ΔAUROC = 0.128 for CRS vs. 0.234 for Delphi-2M), likely because BMI and smoking were included as predictors in Delphi-2M but not in CRS. Since the Delphi-2M model parameters were not publicly available, we performed an indirect comparison by evaluating CRS in held-out individuals matched to the testing population described in Shmatko et al., using the same 2021–2022 test window and a one-year gap between predictor diagnoses and outcomes (Methods). In principle, nonlinear models are expected to outperform linear models given sufficient sample sizes and optimal training ^31^; the superior performance of CRS observed here suggests that, for comorbidity risk prediction, increasing sample sizes currently outweighs adding model complexity.

**Fig. 2:**
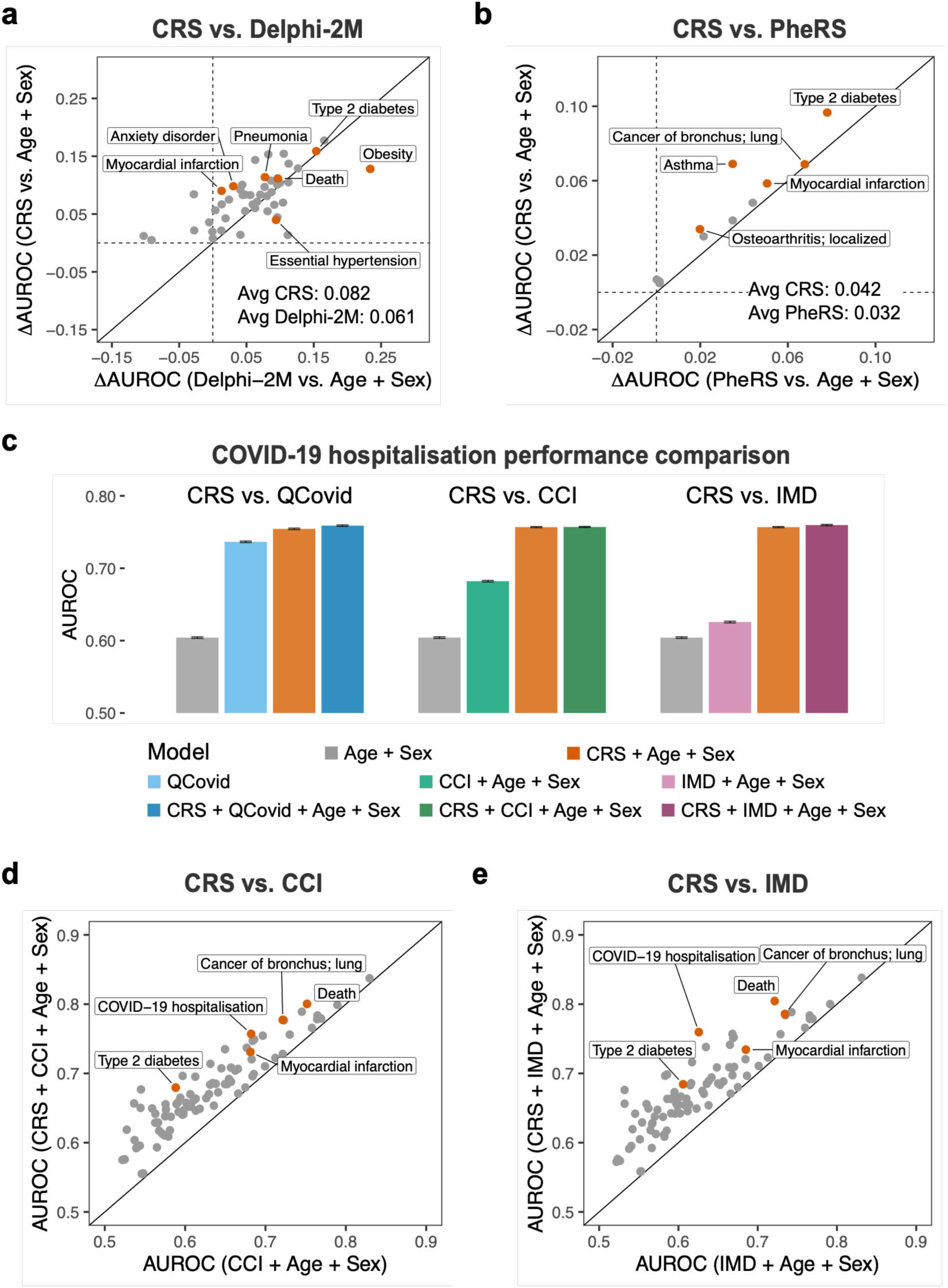
CRSs outperform state-of-the-art EHR risk models. **a.** Improvement in AUROC (ΔAUROC) by CRS over the age-and-sex baseline (*y*-axis) vs. ΔAUROC by Delphi-2M over the age-and-sex baseline (*x*-axis). Due to unavailability of Delphi-2M model parameters, an indirect comparison was performed by evaluating CRS in held-out individuals matched to the testing population in Shmatko et al. for 49 overlapping outcomes. Average ΔAUROC across outcomes for CRS and Delphi-2M are reported in text. **b.** ΔAUROC by CRS over the age-and-sex baseline (*y*-axis) vs. ΔAUROC by PheRS over the age-and-sex baseline (*x*-axis) for 11 overlapping outcomes. Average ΔAUROC across outcomes for CRS and PheRS are reported in text. **c.** Improvement by CRS over existing clinical risk models in predicting COVID-19 hospitalisation. For each competing model (QCovid, CCI, and IMD), AUROC (*y*-axis) is displayed for (1) age-and-sex baseline, (2) competing model, (3) CRS combined with age and sex, and (4) CRS combined with competing model, age, and sex. Error bars represent 95% CI around AUROC. **d.** Improvement by CRS over CCI for all 88 outcomes. AUROC for the model including CRS, CCI, age, and sex (*y*-axis) vs. AUROC for the model including only CCI, age, and sex (*x*-axis). **e.** Improvement by CRS over IMD for all 88 outcomes. AUROC for the model including CRS, IMD, age, and sex (*y*-axis) vs. AUROC for the model including only IMD, age, and sex (*x*-axis). Source data are reported in Table S13 (Panel a), Table S14 (Panel b), Table S15 (Panel c), Table S16 (Panel d), and Table S17 (Panel e).

We next compared CRS with PheRS ^3^, a linear model trained with regularisation (elastic net) in the UK Biobank (*N* = 0.5 million; Fig. 2b). Across the 11 overlapping outcomes, the average ΔAUROC for CRS (0.042 ± 0.000) was 30% higher than that for PheRS (0.032 ± 0.000). The much larger training sample used for CRS than that of PheRS also enabled prediction of many less prevalent outcomes (88 outcomes for CRS vs. 13 for PheRS). In addition, CRS can estimate the effects sizes of highly correlated predictors on future disease outcomes, whereas effect estimates from regularised regression models such as PheRS are primarily useful for prediction rather than statistical inference. For example, consistent with prior studies on risk factors for T2D ^32,33^, CRS estimated the log odds ratio of lipid metabolism disorders on T2D to be 0.245 ± 0.006, conditional on all other prior comorbidities, whereas PheRS shrank the effect of lipid metabolism disorders on T2D below zero (Fig. S7).

We tested the added value of CRS over existing clinical risk models for COVID-19 hospitalisation (Fig. 2c). CRS outperformed QCovid (AUROC = 0.754 ± 0.001 for CRS + age + sex vs. 0.737 ± 0.001 for QCovid), a state-of-the-art model for predicting COVID-19 hospitalisation ^4,10^. Adding QCovid to CRS only slightly improved predictive performance over CRS alone (AUROC = 0.759 for CRS + QCovid + age + sex), despite QCovid including many predictors not directly used in CRS, such as medication usage and organ transplant history ^10^. This suggests that CRS captured the effects of a wide range of risk factors using prior comorbidities alone. Similarly, CRS outperformed the Charlson Comorbidity Index (CCI ^20^; AUROC = 0.682 for CCI + age + sex) and the Index of Multiple Deprivation (IMD ^34^; AUROC = 0.626 for IMD + age + sex). Combining CCI or IMD with CRS did not improve predictive accuracy over CRS alone, suggesting that prior comorbidities captured much of the information contained in existing clinical comorbidity indices and socioeconomic indices.

Lastly, CRS added significant predictive value beyond existing clinical comorbidity and socioeconomic indices across all outcomes (Fig. 2d-e). Comorbidity burden is often associated with lower socioeconomic status ^35^ and commonly summarised using weighted comorbidity indices to inform clinical decision-making ^36,37^. We found that CRS significantly improved predictive performance beyond both CCI (average AUROC = 0.678 for CRS + CCI + age + sex vs. 0.629 for CCI + age + sex) and IMD (average AUROC = 0.680 for CRS + IMD + age + sex vs. 0.627 for IMD + age + sex). However, adding CCI or IMD to CRS did not improve predictive accuracy over CRS alone for most outcomes (Fig. S8). Taken together, these results suggest that CRS has the potential to substantially improve the clinical utility of prior comorbidities for disease risk prediction.

### CRSs retain predictive power across ethnic groups

CRS trained in White British individuals retained high predictive accuracy in held-out samples from multiple self-reported ethnic groups (Fig. 3a), in contrast to genetics-based PRSs, which have poor cross-ancestry transferability ^38^. Compared to held-out White British individuals (average AUROC = 0.678), AUROC was on average retained at 98.1% in Asian individuals, 97.3% in Black individuals, 98.6% in Mixed individuals, and 99.9% in Irish/Other White individuals. For example, the CRS for myocardial infarction achieved an AUROC of 0.744 in Asian individuals and 0.745 in Black individuals, compared to 0.732 in White British individuals. In addition, the CRS for death achieved an AUROC of 0.801 in Asian individuals and 0.762 in Black individuals, compared to 0.798 in White British individuals. The consistent performance of CRS across self-reported ethnic groups suggests that, for most outcome diseases, the effects of prior comorbidities on future disease risk are robust to differences in genetic backgrounds ^3,39^.

**Fig. 3:**
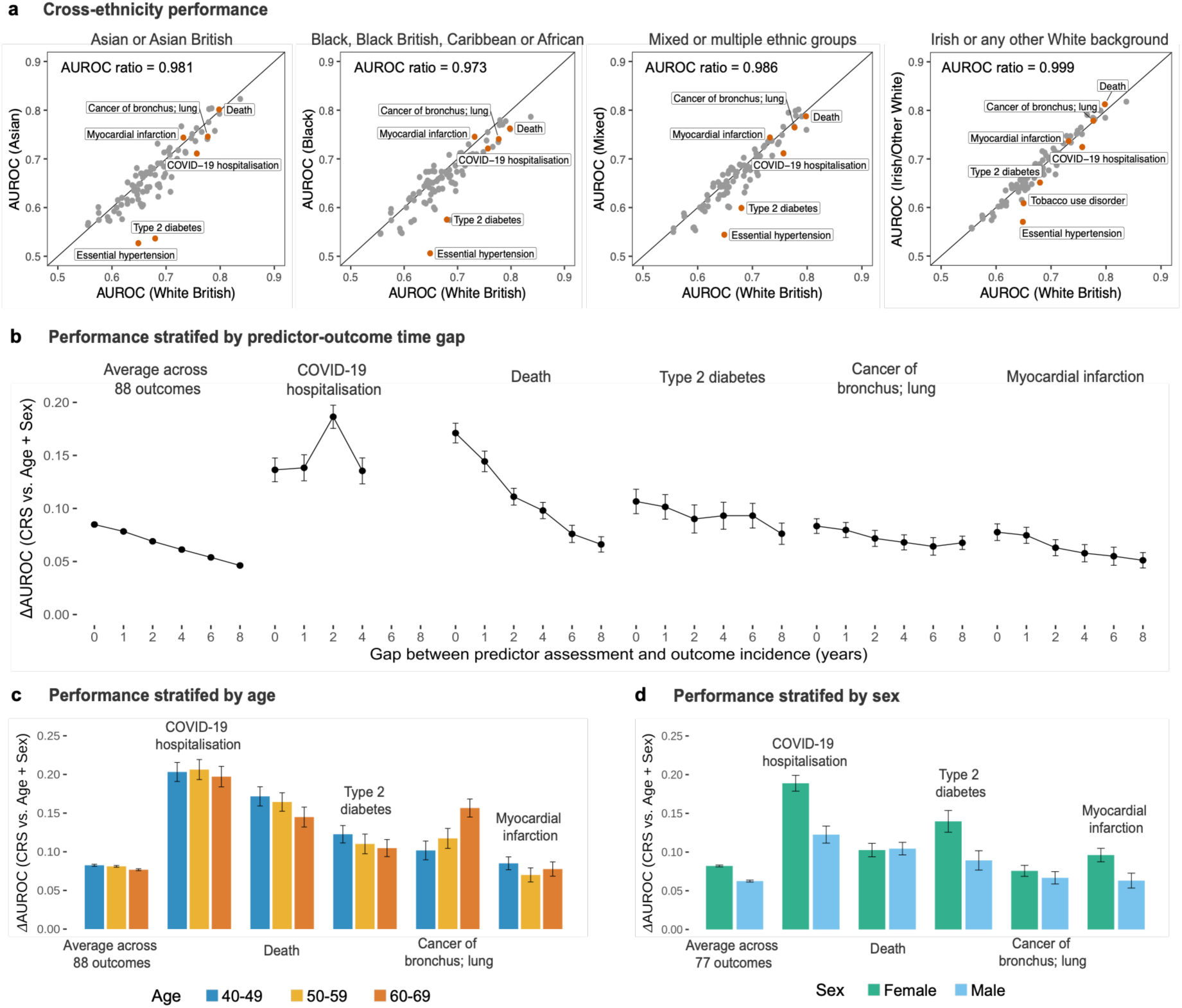
CRSs retain predictive power across ethnicity, longitudinal, and demographic strata. **a**. Cross-ethnicity testing performance of CRS trained in White British individuals. AUROC assessed in held-out Asian, Black, Mixed, and Irish/Other White ethnicity groups (*y*-axis) are separately compared against AUROC assessed in held-out White British group (*x*-axis). All 88 outcomes are shown, with example outcomes labelled and highlighted in red points. In each comparison, the slope of a linear regression through the origin is reported in text. **b.** CRS performance stratified by predictor-outcome time gap. Improvement in AUROC by CRS over the age-and-sex baseline (ΔAUROC; *y*-axis) vs. gap between predictor assessment and outcome incidence in years (*x*-axis) is shown for example outcomes, with the default gap being 3 months. Error bars represent 95% CI around ΔAUROC (very narrow for the average across 88 outcomes). **c.** ΔAUROC stratified by age groups for example outcomes. Error bars represent 95% CI around ΔAUROC. **d.** ΔAUROC stratified by sex for example outcomes. Average ΔAUROC was computed across 77 non-sex-specific outcomes. Error bars represent 95% CI around ΔAUROC. Source data are reported in Table S18 (Panel a) and Table S7 (Panel b-d).

Despite the consistent predictive accuracy of CRS for most outcomes, CRS for essential hypertension and T2D showed poor transferability in self-reported Asian and Black individuals. This likely reflects differences in the features of disease development across ethnic groups as reported in previous studies. For example, although T2D is strongly associated with obesity in European populations, East Asian and South Asian populations tend to develop T2D at lower BMI and younger ages ^40,41^. In addition, standard metabolic syndrome criteria often fail to identify T2D cases in Black populations ^42^.

We performed two secondary analyses. First, we compared CRS performance using improvement in AUROC vs. the age-and-sex baseline across the four self-reported ethnic groups and obtained similar conclusions (Fig. S9). Second, we evaluated transferability using different performance metrics and reached similar conclusions across four other metrics (Fig. S10). Together, these results show that CRS accurately predicts disease outcomes across multiple self-reported ethnic groups.

### CRSs retain predictive power across longitudinal and demographic strata

The effects of prior comorbidities on future disease outcomes persisted for years after their diagnosis. We evaluated the predictive performance of CRS for outcomes occurring after varying gaps from predictor diagnoses (Fig. 3b, Methods). Averaged across 88 outcomes, CRS retained 54.5% predictive power after a gap of 8 years and 3 months (ΔAUROC = 0.046), relative to its performance after 3 months (ΔAUROC = 0.085). For lung cancer and myocardial infarction, CRS retained 81.1% and 65.9% of its predictive power, respectively, after a gap of 8 years and 3 months, compared to its performance after a 3-month gap. CRS based on prior diseases recorded before 1 October 2019 showed consistent predictive accuracy for COVID-19 hospitalisation between 1 January 2020 and 31 December 2024, despite the emergence of different SARS-CoV-2 variants over this period. Together, these results show that CRS retains its power for predicting disease outcomes several years after prior comorbidities were diagnosed, demonstrating the long-term effects of prior comorbidities in addition to more immediate effects on future health outcomes.

We also evaluated CRS performance in testing samples stratified by age and sex. CRS showed similar performance across three age groups, with an average ΔAUROC (vs. the age-and-sex baseline) of 0.083 in patients aged 40–49 years, 0.081 in patients aged 50–59 years, and 0.077 in patients aged 60–69 years (Fig. 3c). CRS was more predictive in older individuals for diseases such as lung cancer, which may reflect age-dependent disease subtypes with distinct comorbidity architectures ^39,43,44^. CRS was also more predictive in females (average ΔAUROC = 0.082) than in males (average ΔAUROC = 0.062, Fig. 3d), potentially due to predictor comorbidities that disproportionately affect females. For example, anxiety disorders, almost twice likely to be diagnosed in females vs. males ^45^, were among the top predictors for many outcomes including myocardial infarction (Table S5). Similarly, multiple sclerosis, another disease that disproportionately affects females ^46^, was a top predictor of COVID-19 hospitalisation. To test whether these conclusions depended on the metric used, we repeated the stratified analyses using C-index, liability-scale *R*^2^, observed-scale *R*^2^, and AUPRC (Table S7) and obtained similar results.

We conclude that CRS captures the long-term effects of prior comorbidity on future disease outcomes and maintains predictive performance across age and sex strata.

### CRSs reveal the comorbidity architecture of common diseases

CRSs accurately estimate the effects of prior comorbidities on disease outcomes conditional on other comorbidities. We defined *comorbidity architecture* of a disease outcome as the distribution of effect sizes of prior comorbidities on that outcome (Methods). We used the effective number of independently predictive diseases, *M̂*_*e*_(estimated using 4^th^ moments; see Methods), to characterise comorbidity architecture ^47^ (Fig. 4a). Intuitively, disease outcomes determined by a few strong comorbidities will have small *M̂*_*e*_’s, whereas outcomes influenced by many prior comorbidities with smaller effects will have large *M̂*_*e*_’s. We simulated comorbidity data using empirical comorbidity prevalences and correlations and found that *M̂*_*e*_ accurately estimated the effective number of independently predictive comorbidities (Methods; Fig. S11). Comorbidity architecture varied substantially across disease outcomes in the empirical data. For example, T2D (*M̂*_*e*_ = 12.6), lung cancer (*M̂*_*e*_ = 15.1), and myocardial infarction (*M̂*_*e*_ = 25.2) were dominated by a few strong predictors with large effects, whereas COVID-19 hospitalisation (*M̂*_*e*_ = 61.2) and death (*M̂*_*e*_ = 52.1) reflected contributions from many weak predictors with small effects. Averaged across 88 disease outcomes, 42 prior comorbidities were independently predictive of future disease outcomes (average *M̂*_*e*_ = 41.9 ± 2.3). *M̂*_*e*_ quantifies the distribution of effect sizes, which is different from the number of prior comorbidities with significant effect sizes (average number of significant predictors per outcome = 101.4 ± 3.3; Table S5).

**Fig. 4:**
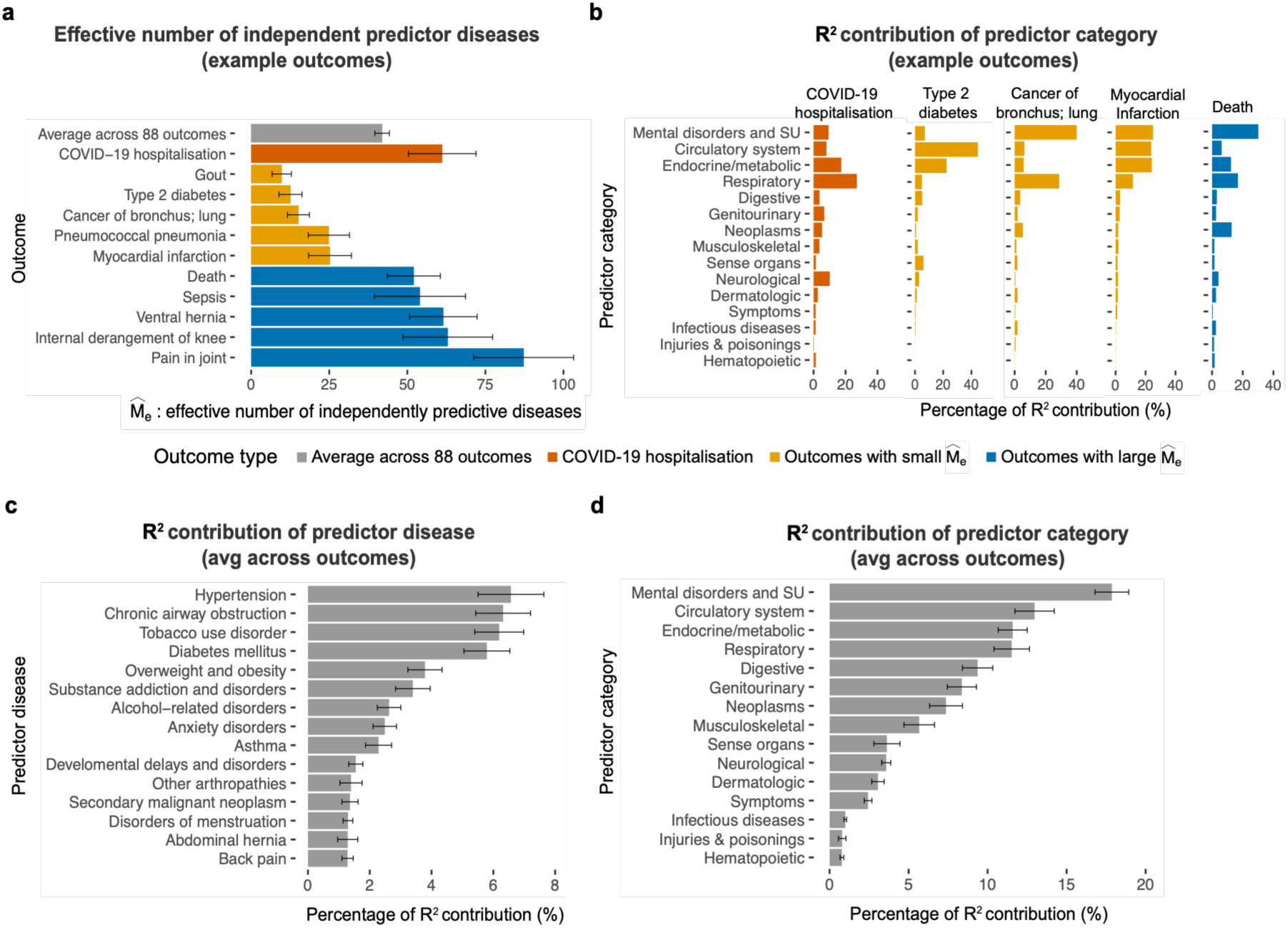
CRSs reveal the comorbidity architecture of common diseases. **a.** Effective number of independently predictive diseases, *M̂*_*e*_ (*x*-axis), for example outcomes (*y*-axis). Error bars represent standard errors around the point estimates *M̂*_*e*_. **b.** Percentage of CRS *R*^2^ contribution (*x*-axis) from predictor categories as defined by Phecode categories (*y*-axis) for example outcomes. **c.** Percentage of CRS *R*^2^ contribution (*x*-axis) from top 15 predictor diseases (*y*-axis) averaged across 88 outcomes, ordered by magnitude of contribution. Error bars represent standard errors around the mean percentage contribution, averaged across outcomes. **d.** Percentage of CRS *R*^2^ contribution (*x*-axis) from each predictor category (*y*-axis) averaged across 88 outcomes, ordered by magnitude of contribution. Error bars represent standard errors around the mean percentage contribution, averaged across outcomes. Abbreviations: SU, substance use. Source data are reported in Table S19 (Panel a), Table S20 (Panel b), Table S21 (Panel c), and Table S22 (Panel d).

CRS prioritised prior comorbidities and Phecode categories with large effects on health outcomes over other correlated comorbidities. We quantified the relative contribution of each prior comorbidity and category using its contribution to the *R*^2^ of the CRS (Methods). CRS for disease outcomes with a small *M̂*_*e*_was dominated by a few prior comorbidities (Fig. 4b). For example, CRS for lung cancer (*M̂*_*e*_ = 15.1) was dominated by “mental disorders and substance use” category and “respiratory” category, which jointly contributed 69% of the predictive *R*^2^ of the lung cancer CRS. Within these two categories, tobacco use disorder and chronic airway obstruction contributed 60% of the predictive *R*^2^. By contrast, CRS for disease outcomes with a large *M̂*_*e*_ had effect sizes distributed across many comorbidities with moderate to small effects. For example, CRS for COVID-19 hospitalisation (*M̂*_*e*_ = 61.2) had 7 predictor categories each contributing more than 5% of the *R*^2^. No single prior comorbidity contributed more than 12% of the predictive *R*^2^for the COVID-19 hospitalisation CRS. The prior comorbidities prioritised by CRS were consistent with previous studies, such as smoking for lung cancer ^48^, COPD for severe COVID-19 ^49^, and hypertension, diabetes, and dyslipidaemia for myocardial infarction ^50^.

CRS identified health-critical comorbidities that elevated risk across many disease outcomes (Fig. 4c). Averaged across 88 disease outcomes, hypertension (average CRS *R*^2^ contribution = 6.6 ± 1.1%), chronic airway obstruction (6.3 ± 0.9%), tobacco use disorder (6.2 ± 0.8%), and diabetes (5.8 ± 0.7%) were the top health-critical comorbidities. Aggregating comorbidities in each disease category, we found that the “mental disorder and substance use” category had the highest contribution to CRS (average *R*^2^ contribution = 17.9 ± 1.1% across 88 outcomes; Fig. 4d), which included health-critical comorbidities such as tobacco use disorder, substance addiction and disorders, alcohol-related disorders, and anxiety disorders. “Circulatory system” (average *R*^2^ contribution = 13.0 ± 1.2%), “endocrine/metabolic” (11.6 ± 0.9%), and “respiratory” (11.5 ± 1.1%) categories also had large contributions to CRS. These health-critical comorbidities and categories contributed substantially to the risk of many disease outcomes when conditioned on all other comorbidities, which may inform clinical decision-making to prevent adverse outcomes in individuals with multiple long-term conditions.

### CRSs identify individuals at high absolute risk

CRSs were trained on individuals aged 40–69 from nearly the entire population of England, making them well-suited to population-level disease screening. For common disease outcomes, CRSs prioritise large numbers of high-risk individuals (Fig. 5a). For example, the CRS for myocardial infarction identified 316,955 testing individuals in 2015 within top 5% of the score, of whom 41,870 developed myocardial infarction between 2015 and 2024, accounting for 19.4% of all cases in the testing population. Similarly, the CRS for type 2 diabetes identified 297,420 testing individuals in 2015 within top 5% of the score, of whom 68,670 developed T2D between 2015 and 2024, accounting for 13.2% of all cases. The top 5% risk group predicted by CRS had 73% higher disease incidence on average than the top 5% predicted by an age-and-sex baseline model (Table S8). Furthermore, the CRS was appropriately calibrated for predicting disease risk among the English population (Fig. 5b; calibrated CRS coefficients in Table S9). The observed risk in the top 5% of CRS scores was slightly lower than predicted, which may reflect preventive treatment initiation and non-random censoring among high-risk individuals.

**Fig. 5:**
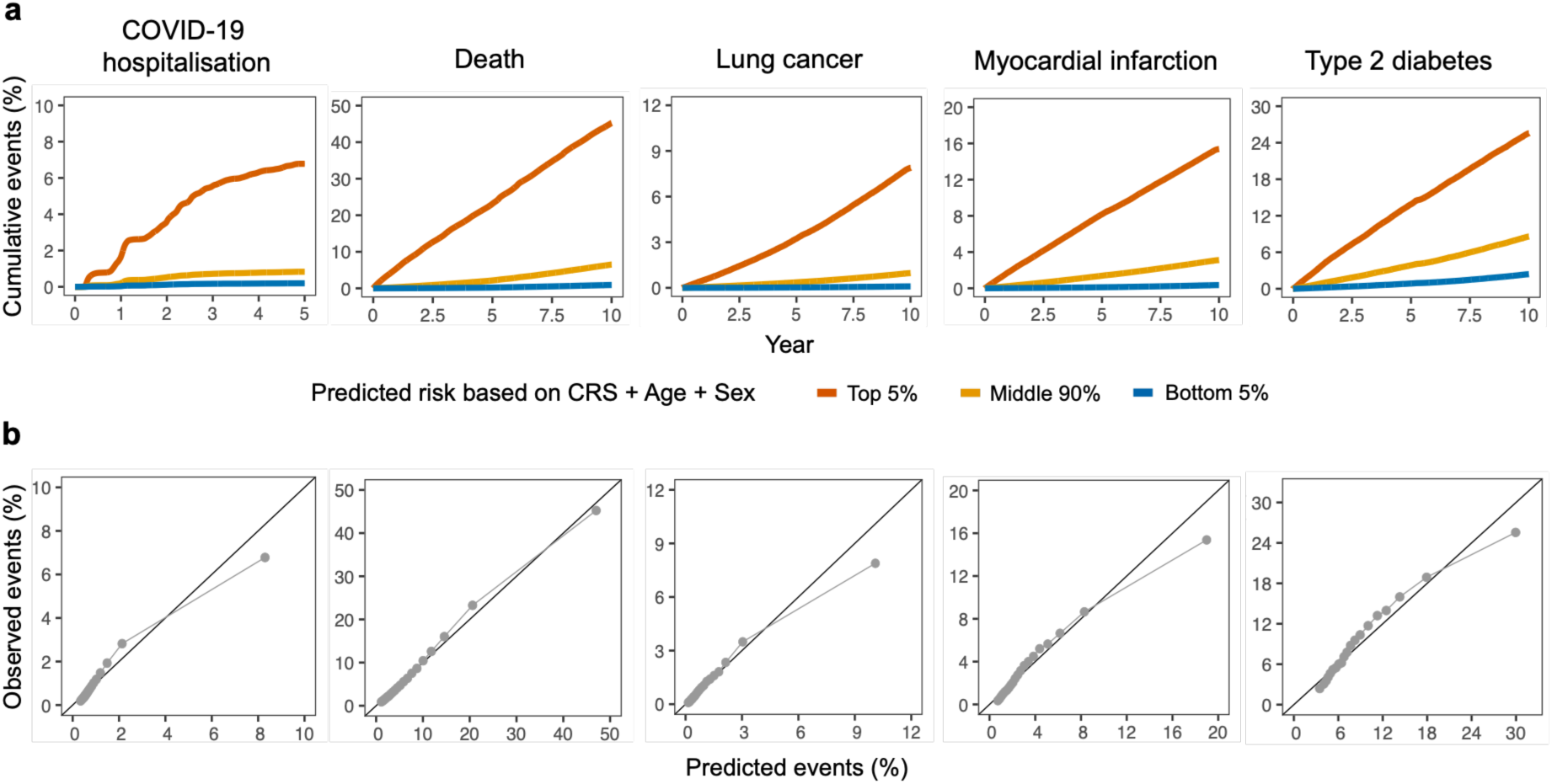
CRSs identify individuals at high absolute risk with good calibration. **a.** Observed cumulative event rate (*y*-axis) vs. year since follow-up (*x*-axis) for example outcomes, stratified by top 5%, middle 90%, and bottom 5% quantiles of predicted risk based on the model including CRS, age, and sex among the testing population. 95% CI around cumulative event rate is provided but too narrow to be visible. **b.** Calibration of CRS for example outcomes. For each outcome, the testing population was divided into 20 equal quantiles based on predicted risk from the model including CRS, age, and sex. Observed event rate within each quantile (*y*-axis) is compared against average predicted event rate within each quantile (*x*-axis). Source data are reported in Table S23-S27 (Panel a) and Table S28 (Panel b).

### CRSs distinguish independently predictive comorbidities from indirect associations

Marginal comorbidity-outcome associations can reflect indirect effects when mediating comorbidities are not included as covariates. By contrast, CRS included nearly all prior diagnoses (outside the target disease Phecode category) as covariates during training. Therefore, its effect estimates are conditioned on other comorbidities and quantify independently predictive effects. We highlight two examples in which CRS elucidated marginal effects that were mediated by other comorbidities. First, we found that tobacco use disorder was a strong independent predictor of sepsis but not of COVID-19 hospitalisation (Fig. 6a). Sepsis and COVID-19 hospitalisation had similar CRS effect sizes (Pearson’s *r* = 0.70), consistent with previous studies reporting an association between COVID-19 hospitalisation and sepsis ^51^. However, tobacco use disorder strongly predicted sepsis (CRS effect size = 0.130 ± 0.003) but not COVID-19 hospitalisation (CRS effect size = −0.017 ± 0.004) when conditioning on other predictors, consistent with existing studies reporting weak negative association between smoking and COVID-19-related deaths after adjusting for comorbidities ^52^. We performed a secondary analysis by estimating the effect size of tobacco use disorder on COVID-19 hospitalisation while conditioning on different numbers of other comorbidities (Fig. 6a, right panel). The marginal effect size of tobacco use disorder on COVID-19 hospitalisation was 0.182 ± 0.003 and dropped to 0.003 ± 0.004 when the top 50 common comorbidities were included as covariates. Including chronic airway obstruction or pneumonia as covariates caused the greatest attenuation (Fig. S12). Our results support distinct roles of smoking in affecting COVID-19 and sepsis: while smoking mainly increases severe COVID-19 risk through respiratory diseases ^53,54^, it may confer additional independent risk for sepsis through pathways not captured by respiratory diseases ^55^.

**Fig. 6:**
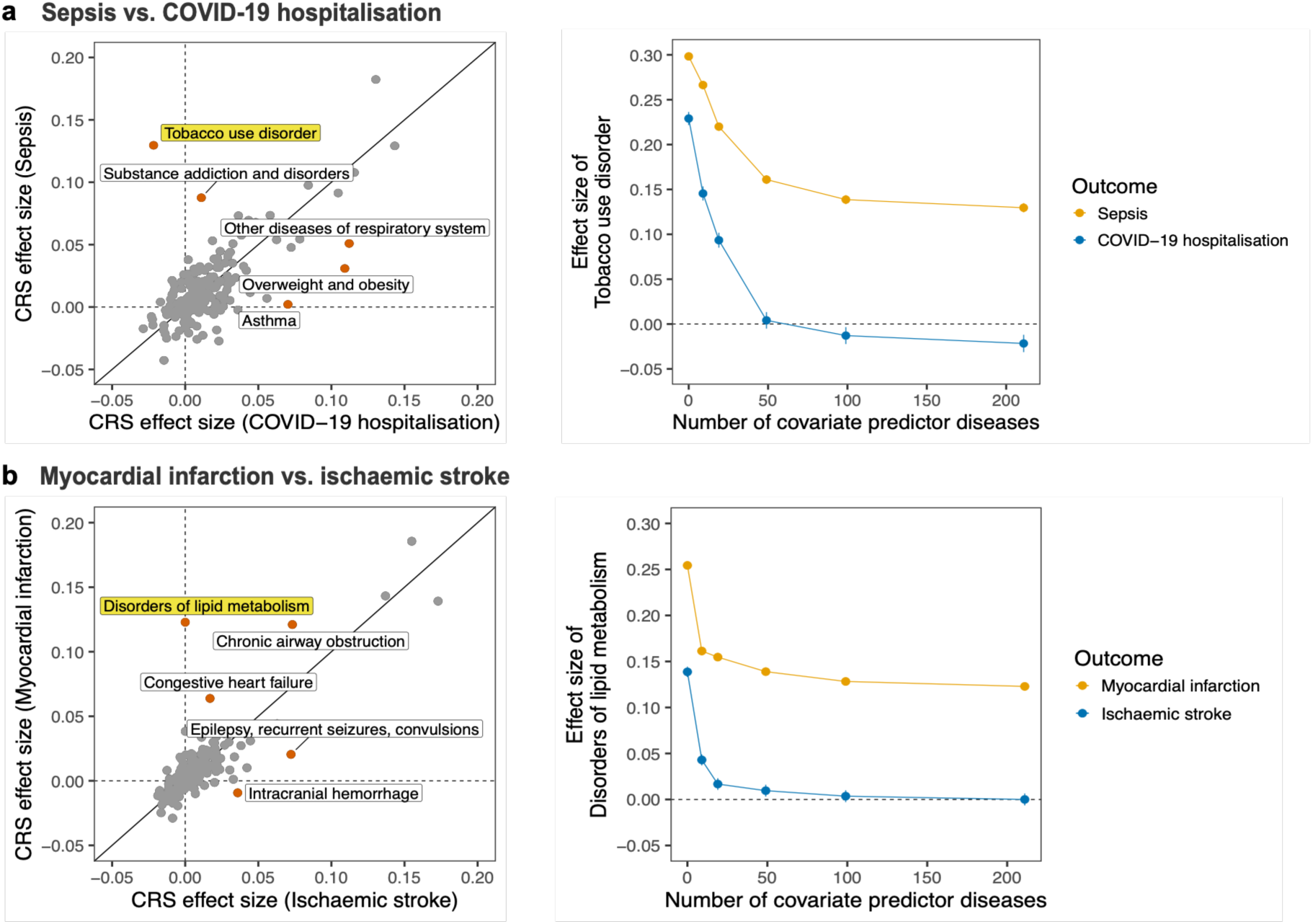
CRSs distinguish independently predictive comorbidities from indirect associations. **a.** Distinct predictive roles of tobacco use disorder in sepsis vs. COVID-19 hospitalisation. Left, CRS effect sizes (defined as standardised log odds ratio of predictors) for sepsis (*y*-axis) vs. CRS effect sizes for COVID-19 hospitalisation (*x*-axis). The top 5 predictors with largest differences in effect size between outcomes are labelled and highlighted in red points; tobacco use disorder is further highlighted in yellow label. Right, effect size of tobacco use disorder (*y*-axis) vs. number of covariate predictor diseases included in CRS (*x*-axis) grouped by outcome. **b.** Distinct predictive roles of lipid metabolism disorders in myocardial infarction vs. ischaemic stroke. Left, CRS effect sizes for myocardial infarction (*y*-axis) vs. CRS effect sizes for ischaemic stroke (*x*-axis). The top 5 predictors with largest differences in effect size between outcomes are labelled and highlighted in red points; disorders of lipid metabolism is further highlighted in yellow label. Right, effect size of lipid metabolism disorders (*y*-axis) vs. number of covariate predictor diseases included in CRS (*x*-axis) grouped by outcome. Source data are reported in Table S29 (Panel a and b, left) and Table S30 (Panel a and b, right).

Similarly, we found that lipid metabolism disorder was independently predictive of myocardial infarction but not of ischaemic stroke (Fig. 6b). Despite similar CRS effect sizes for myocardial infarction and ischaemic stroke (Pearson’s *r* = 0.83), lipid disorder was an independent predictor of myocardial infarction (CRS effect size = 0.123 ± 0.002), but not of ischaemic stroke (CRS effect size = 1e-05 ± 0.003) when conditioning on other comorbidities. The marginal effect of lipid disorder on ischaemic stroke was 0.139 ± 0.003 and dropped to 0.010 ± 0.003 when the top 50 common comorbidities were included as covariates. Including ischaemic heart disease, hypertension, and diabetes caused the greatest attenuation (Fig. S12). This result was consistent with previous studies reporting weaker signals of lipid disorder on ischaemic stroke than on coronary heart disease ^56,57^. While lipid disorder may have a direct effect on myocardial infarction through coronary atherosclerosis, its effect on ischaemic stroke may be largely mediated by other comorbidities capturing vascular atherosclerotic burden. Overall, we demonstrated CRS’s ability to distinguish independently predictive comorbidities from indirect associations, providing a valuable resource for further investigation of disease mechanisms.

## Discussion

The COVID-19 pandemic highlights the vulnerability of individuals living with multiple long-term conditions, who were more susceptible to severe COVID-19 illness and other adverse outcomes. In this study, we developed CRSs to predict COVID-19 hospitalisation and 87 other disease outcomes based on past disease diagnoses, leveraging large-scale nationwide EHR data. CRSs were trained at close to saturated sample size and provide a resource quantifying the effect of 212 common prior diseases on the 88 outcomes. In predicting COVID-19 hospitalisation, past disease diagnoses alone performed better than state-of-the-art clinical COVID-19 models. CRSs were powerful at identifying individuals with high disease risks across different ethnic groups and outperformed state-of-the-art AI models and established clinical risk scores. Together, CRSs offer high predictive power, interpretability, and generalisability, with the potential to improve clinical risk prediction.

CRSs provide both clinical impact and insights into disease aetiology. Averaged across disease outcomes, CRSs identified 5% of the population with 3.4-fold higher disease risk. Such a level of risk enrichment is broadly in line with established guidelines for clinical implementation of risk scores (e.g., eMERGE consortium ^58^). The cross-ethnicity transferability of CRSs further supports their potential clinical utility in ethnically diverse populations. On the other hand, CRS effect sizes highlighted key comorbidities that independently predict future disease risks, many of them replicating established risk factors. The consistency between CRS correlation and genetic correlation across outcomes validates the ability of CRSs in capturing molecular mechanisms underlying the comorbidity-outcome associations. Outcomes with similar CRS profiles can thus be further investigated for shared biological pathways.

Notably, the logistic regression-based CRSs outperformed Delphi-2M, trained using GPT models from UK Biobank data, in predicting most outcomes. Previous work on polygenic scores and clinical risk prediction ^59–61^ revealed similar patterns where deep learning models failed to outperform simpler regression models in trait and disease prediction with biobank-scale sample sizes (∼0.5 million). Our results further suggested the advantage of training simpler models with very large-scale data. Given that GPT models are still in early stages of adaptation to EHR data modelling, they will benefit from further optimisation and tailoring to achieve more powerful risk prediction. In addition, the lack of advantage by Delphi-2M may indicate limited nonlinear effects of diagnoses on disease outcomes, as observed for genetic effects on complex traits ^62^. Still, we note that this was an indirect comparison of two methods tested in different cohorts, where the study populations can have systematic differences: while CVD-COVID-UK covers the general population from England, the UK Biobank cohort is known to be healthier than the general population ^63^, often resulting in worse predictive performance. A direct comparison in future studies will provide more concrete conclusions.

Our study had several other limitations. First, as with any EHR-based research, our reported CRS effect sizes and model performance depended on the phenotypic definitions. Here we used Phecodes to define predictor and outcome diseases, as consistent with a range of prior EHR studies performing phenome-wide analyses ^64,65^. These definitions reflect the underlying healthcare-seeking and diagnostic practices rather than clinically assessed disease onset, thus susceptible to bias caused by underdiagnosis or selection of health-aware or sicker patients. Second, for model simplicity, we treated predictor diseases as binary presence/absence and did not account for the age of diagnosis or orders of comorbidity onset. Such information may carry additional predictive power ^1^. Furthermore, while the CRSs were developed based on the population of England and applicable to the healthcare system in England (NHS England), it will be of interest to further evaluate CRS in other healthcare systems. Lastly, while our estimated CRS effect sizes capture predictive power of past diseases on future outcomes, they cannot be interpreted as causal effects since this was an observational study without formal causal inference.

Several new directions are promising based on the current study. As a comprehensive summary of individuals’ prior diagnosis profile, CRS not only provides powerful prediction by itself, but can also be combined with other established risk prediction tools. For example, EHR-based risk scores have been shown to offer additional prediction improvement over genetics-based PRS ^3^. With the increasing availability of linkage between EHR data and other data modalities, such as genetics, biomarkers, and multi-omics data ^66–68^, it will be interesting to further evaluate the additive benefit provided by CRSs over other data types. On the other hand, while we have developed CRSs using simpler logistic regression models, novel methodologies can be explored to capture more complex information within EHR data. For example, we can incorporate interaction effects across diagnoses as well as the effect of comorbidity onset. A useful direction will be to increase model complexity for better performance without compromising the interpretability or generalisability of the models.

In conclusion, we have demonstrated CRSs as a valuable resource for clinical risk prediction across a wide range of diseases. The powerful prediction of COVID-19 hospitalisation and related diseases provides evidence for future pandemic planning where individuals at high risk based on their comorbidities could be identified for targeted intervention. With a growing focus on early detection and prevention of diseases in healthcare settings worldwide, our work offers an accurate and accessible toolset that benefits risk assessment at a population scale.

## Materials and methods

### Data description

We performed our analyses using electronic health records (EHRs) from the CVD-COVID-UK cohort, an England-wide EHR data resource that enables population-level research on COVID-19 and cardiovascular diseases ^15^. We accessed anonymised, national, linked EHRs for England in NHS England’s (NHSE) Secure Data Environment (SDE) service, made available via the British Heart Foundation Data Science Centre’s CVD-COVID-UK/COVID-IMPACT Consortium. The datasets contained linked individual-level records from primary care, secondary care, death registry, and COVID-19 laboratory test results. To capture disease diagnoses from both primary care and hospital settings, we focused on the combined EHR data from the primary care ^69^ and Hospital Episode Statistics Admitted Patient Care (HES APC) ^70^. As primary care data was only available for individuals alive on 1 January 2020 due to the original CVD-COVID-UK cohort design ^15^, 92.2% of individuals had primary care data available in our study population (see Section “Study design”).

Curation and cleaning of EHR data were performed following the Health Data Science Team Documentation ^71^ by the British Heart Foundation Data Science Centre. Compliant with the NHS SDE guidance on data outputs, all reported counts (e.g., sample sizes) in this manuscript were rounded to the nearest multiple of 5.

### Study design

We developed comorbidity risk scores (CRSs) to predict the risk of COVID-19 hospitalisation and 87 other incident disease outcomes among adult patients aged 40-69 years old in England, using longitudinal disease diagnoses in the combined EHR data from primary care and HES APC in the CVD-COVID-UK cohort. Fig. S1a shows a schematic overview of the study design. Patients within the target age range on a fixed index date (1 January 2020 for COVID-19 hospitalisation; 1 January 2015 for other disease outcomes) were followed up for up to 5 (COVID-19 outcome) or 10 years (other outcomes) to assess the incidence of outcome diseases. Prior diagnoses for 212 common diseases 3 months before the index date were used as predictors; the gap of 3 months was introduced to avoid predicting incident events recorded soon after prior diagnoses, which may be part of the outcome diagnostic process. For each outcome and predictor disease, only the earliest recorded diagnosis per individual from either primary care or HES APC was used. Criteria for selection of outcomes and predictors are described in Section “Outcomes and predictors in CRS”.

CRS was first trained in self-reported White British individuals born in odd years and tested in White British individuals born in even years. To evaluate cross-ethnicity performance, CRS was also tested in all individuals, regardless of birth year, who self-reported as Asian, Black, Mixed and Irish/Other White (see Section “Cross-ethnicity and stratified evaluation”). To fully employ the national-level sample size of the CVD-COVID-UK cohort, we also trained CRS in all White British individuals and released the corresponding effect size estimates (Table S3).

The inclusion criteria for the study population were as follows: (1) aged 40-69 years old on the *index date*, defined as 1 January 2020 for COVID-19 hospitalisation outcome, and 1 January 2015 for other disease outcomes; (2) had ≥ 1 diagnosis of ≥ 1 predictor disease 3 months before the index date in either primary care or HES APC; (3) alive and had ≥ 1 day of follow-up from the index date, where the *follow-up period* was defined as the period from index to the earliest of 31 December 2024 or death. Individuals with the following criteria were excluded from the study population: (1) for each disease outcome, individuals who had any diagnosis of the outcome before the index date; (2) had missing data in age, sex, or ethnicity; (3) had one-time-use ID as their unique patient identifier (Person_ID) ^72^. The overview of sample selection is shown in Fig. S1b-c.

For each disease outcome, the study population was further selected into case and control groups. In training cohorts, cases were defined as individuals who had ≥ 1 diagnosis of the target disease outcome during the follow-up period, and controls were defined as individuals who had follow-up till 31 December 2024 with no diagnosis of the target outcome during the follow-up period. For a balance between statistical power and computational efficiency, a subset of all eligible controls was randomly selected with a control-case ratio up to 4:1, for the training of CRS models. To account for individuals whose follow-up ended before 31 December 2024 without experiencing the outcome (i.e., censored before the full 5 or 10-year period ended), the inverse probability of censoring weighting (IPCW) method was used in model fitting of CRS (see Section “Construction of CRSs” for details).

### Outcomes and predictors in CRS

We constructed CRSs for 88 disease outcomes, including COVID-19 hospitalisation, all-cause mortality (death), and 86 other common diseases. COVID-19 hospitalisation was identified during 1 January 2020 – 31 December 2024 via two sources: (1) presence of ICD-10 code for confirmed COVID-19 (U07.1) as primary diagnosis in HES APC, or (2) confirmed COVID-19 diagnosis in the UKHSA Severe Acute Respiratory Infection (SARI) Watch system (formerly COVID-19 Hospitalisation in England Surveillance System (CHESS)) ^73^. COVID-19 hospitalisation records from both sources were curated from previous studies ^74^ and used to identify the outcome cases.

Aside from death, the 86 other disease outcomes were defined based on Phecode 1.2 ^64^ and selected based on sufficient case numbers and high risk for hospitalisation, using a data-driven approach. First, ICD-10 codes in HES APC and SNOMED codes in primary care data were mapped to Phecodes using Phecode Map 1.2 ^75^: ICD-10 codes were directly mapped to Phecodes, while SNOMED codes were first mapped to ICD-10 codes according to the technical guide by SNOMED International ^76^ before being mapped to Phecodes. Only ICD-10 codes starting with letters A-N were used as they correspond to disease codes. Next, we selected Phecodes that satisfied the following conditions: (1) ≥ 150,000 incident cases during 1 January 2015 – 31 December 2024, among individuals aged 40-69 on 1 January 2015 in primary care + HES APC; and (2) belonged to the top 10 common Phecodes, or accounted for ≥ 50% individuals’ primary reason for hospital visit (i.e., as primary diagnosis) among individuals who had ≥ 1 target Phecode diagnosis in HES APC, or featured in prior Phecode-based disease prediction studies ^3^. The final list of 88 disease outcomes along with corresponding Phecodes, definitions, and training case numbers is provided in Table S1.

Predictor diseases were similarly defined based on Phecode 1.2 and selected based on prevalence. ICD-10 and SNOMED codes in HES APC and primary care were first mapped to Phecodes, as described before. Phecodes satisfying the following condition were selected as candidates for predictor diseases: ≥ 0.2% lifetime prevalence as of 1 January 2015, among individuals aged 40-69 on 1 January 2015 in primary care + HES APC. To reduce correlations across predictors in CRS models, we further aggregated the 391 candidate Phecodes into 212 parent-level Phecodes. For example, essential hypertension (Phecode 401.1) was aggregated to hypertension (Phecode 401) ^64^. Such an approach ensured that different predictors capture as distinct comorbidities as possible. Lastly, for each disease outcome, we excluded predictors sharing the same parent Phecode as the outcome to avoid including predictor diseases too closely related to the target outcome. For example, prior ischaemic heart disease (Phecode 411) was excluded from the CRS predicting myocardial infarction (Phecode 411.2). The final list of 212 predictor diseases along with corresponding Phecodes, definitions, and prevalence is provided in Table S2.

### Construction of CRSs

For each disease outcome, the CRS was constructed as a weighted linear combination of predictor disease diagnoses. Specifically, for outcome *i*, assuming a total of *M*_*i*_ predictor diseases, the CRS was constructed as follows:

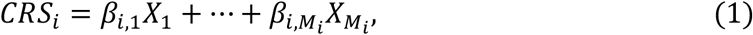

where 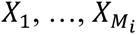 are binary diagnoses of predictor diseases, and the weights 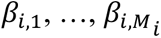 are obtained from the following logistic regression on the training sample:

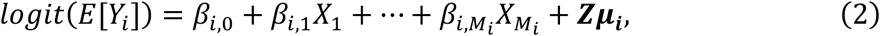

where *Y*_*i*_ is the target disease outcome status within the 5- or 10-year follow-up period, and ***Z*** corresponds to covariates: age at index and binary sex (males vs. females).

Individuals who censored without experiencing the outcome were excluded from model fitting. To account for the potential bias caused by this exclusion (which results in a “healthier” control group), we performed a weighted logistic regression using the IPCW method ^18,77^: non-censored individuals were assigned higher weights if they experienced the outcome later in time, to compensate for the fact that individuals were more likely to censor the longer they were followed up. In addition, sandwich estimators ^78^ were used to produce robust standard errors (SEs) for the predictor effect estimates.

For sex-specific disease outcomes (e.g., prostate cancer in males and female breast cancer in females), the study population was limited to individuals of the applicable sex. Predictor diseases only applicable to the opposite sex were excluded (e.g., prostate cancer was excluded as a predictor when female breast cancer was the outcome), and sex was also excluded as a covariate.

The IPCW-weighted logistic regression approach in model training has several advantages. First, compared to survival models like Cox proportional hazard models, our approach accounts for censoring with fewer assumptions ^18^, while the proportional hazard assumption rarely holds in clinical risk prediction settings ^79,80^. Second, given the large-scale national-wide data, logistic regression is much more efficient computationally than the Cox model which requires estimation of partial likelihood. Lastly, effect size estimates from logistic regression allow us to study the comorbidity architecture underlying CRSs (see Section “Comorbidity architecture of CRSs”), where the metrics being used have been largely derived from regression models rather than survival models.

### Evaluation of CRSs

CRSs were constructed based on the training sample (White British individuals born in odd years) and evaluated in the testing samples, separately tested in held-out White British individuals (born in even years) and other self-reported ethnicity groups. The main evaluation compared the model of interest that included CRS, age, and sex as predictors against the baseline model with age and sex as predictors only.

Predictive performance was evaluated using the following metrics: (1) fold change in cumulative risk for the top 5% of predicted risks: this was captured by the risk ratio comparing top 5% of the predicted risk distribution vs. population average risk, where the observed risks were derived using Kaplan-Meier analyses; (2) area under the receiver operating characteristic curve (AUROC); (3) concordance index (C-index); (4) liability-scale *R*^2^, which captures the proportion of variation in the outcome explained by the predictors on the latent outcome liability scale ^81^; (5) observed-scale *R*^2^, which captures the proportion of variation in the outcome explained by the predictors on the observed binary outcome scale; and (6) area under the precision-recall curve (AUPRC): we reported the difference between AUPRC and the outcome incidence to account for different incidence rates across outcomes.

For each outcome, the fold change in cumulative risk was evaluated in the entire testing population, with the 95% confidence intervals (CIs) around the risk ratio derived from standard errors (SEs) for the Kaplan-Meier estimates of the cumulative risks. We reported the risk ratio estimates for the CRS + Age + Sex model and the baseline Age + Sex model, as well as the fold improvement between the two models.

The other five metrics were evaluated using a Monte Carlo sampling approach for computational efficiency. Consistent with the training samples constructed with a control-case ratio of up to 4:1, for each outcome, we randomly sampled 3,000 cases with a control-case ratio of up to 4:1 (i.e., 15,000 individuals in total) from the entire testing set for 100 replications to assess AUROC, C-index, liability-scale *R*^2^, and observed-scale *R*^2^. To assess AUPRC, which is only meaningful based on realistic outcome incidence, we randomly sampled 1,000 cases with a control-case ratio matching the population-level incidence from the entire testing set for 100 replications. In scenarios where the total case numbers were not sufficiently larger than 3,000 (e.g., in cross-ethnicity or subgroup evaluations), a bootstrapping approach was instead used, where we randomly selected an initial sample of 3,000 cases (or 1,000 cases for AUPRC) with the target case-control ratio before performing bootstrapping on the same sample for 100 replications. For each performance metric, we reported the average metric estimate for the CRS + Age + Sex model and the baseline Age + Sex model, as well as the average difference in performance between the two models, across replications. The 95% CIs were constructed around the metrics based on the 100 replications, assuming a normal distribution of the mean metric estimates.

In the testing samples, we assumed that individuals were controls if they did not experience the outcome any time during their follow-up, including those who censored before the full 5/10-year period. This approach was conservative as the control population was composed of both true controls and potential cases who might develop the outcome if they had complete follow-up, thus making the metrics potentially perform worse. However, since C-index accounts for time to event and censoring when evaluating concordance, it served as a sensitivity metric and we observed high consistency between AUROC and C-index in our results (Fig. S2), justifying our evaluation approach.

### Comparison of CRS effect correlations to phenotypic and genetic correlations

We compared correlations in CRS effect sizes across outcomes (CRS effect correlations) against phenotypic correlations across outcomes. While phenotypic correlations across outcomes are affected by diagnostic error, CRS effect correlations capture the outcomes’ shared comorbidity profile and is not affected by diagnostic error given a sufficient training sample size. To illustrate the effect of diagnostic error, we first assume a liability threshold model for the comorbidity-outcome association, similar to that in genetics literature ^82^. For outcomes *i* and *j*, we assume:

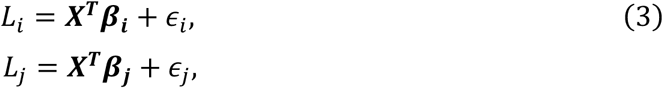

where *L*_*i*_ and *L_j_* are the continuous, unobservable disease liabilities for outcomes *i* and *j*, ***X*** is the data vector containing all predictor disease diagnoses, ***β***_***i***_ and ***β***_***j***_ are random vectors of CRS effect sizes, and 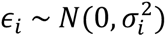 and 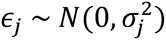 represent independent residuals that contain diagnostic errors.

The phenotypic correlation between outcomes *i* and *j* on the liability scale is thus

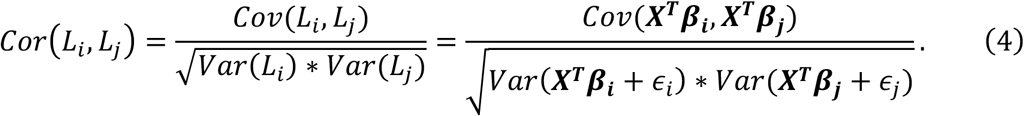

The correlation in comorbidity-predicted disease liability between outcomes *i* and *j* is instead

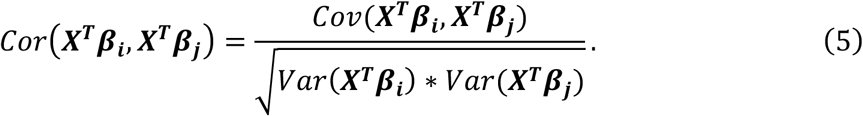

While sharing the same numerator, Eq. (4) has a larger denominator than Eq. (5), due to the additional variance of residual errors captured, resulting in a smaller Eq. (4) than Eq. (5) in magnitude.

If we further assume that the elements of ***β***_***i***_ (and ***β***_***j***_) are independent and the predictor vector ***X*** is standardised with mean 0 and variance 1, it has been shown that ^83^:

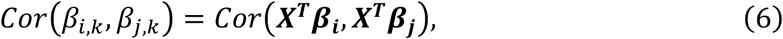

where *β*_*i*,*k*_’s are elements of ***β***_***i***_, and *β_j_*_,*k*_’s are elements of ***β***_***j***_. The left-hand side of Eq. (6) is the correlation across predictor diseases, while the right-hand side is the correlation across individuals. In other words, the CRS effect correlation across predictors between outcomes *i* and *j* is equivalent to the correlation across individuals in comorbidity-predicted disease liability, which is larger in magnitude than liability-scale phenotypic correlation. Given sufficiently large training sample sizes, as in the CVD-COVID-UK cohort, ***β***_***i***_ and ***β***_***j***_ are accurately estimated by the sample estimates 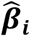 and 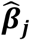, and thus Eq. (6) applies to the estimated CRS effect correlation as well. Furthermore, it has been shown that observed-scale phenotypic correlation (i.e., based on binary disease status) is usually smaller in magnitude than liability-scale phenotypic correlation ^81^. Therefore, CRS effect correlation has the advantage of teasing out comorbidity-related variation from diagnostic error, compared to both observed-scale and liability-scale phenotypic correlations.

To compute the CRS effect correlations, we calculated Pearson’s correlation in standardised CRS effect sizes across all pairs of outcomes, using overlapping predictor diseases for each pair. To compute the observed-scale phenotypic correlations, we calculated Pearson’s correlation in observed binary outcome status across all pairs of outcomes, based on the training samples (*N* = 6.5 million). We then converted the observed-scale phenotypic correlations to liability-scale phenotypic correlations using Monte Carlo simulations under the liability threshold model, as implemented and described in detail in previous studies ^83,84^. For the final comparison of different correlations across outcomes, we removed highly correlated outcomes by filtering out parent and child Phecode pairs, leaving 3,802 eligible outcome pairs in total.

We further compared CRS effect correlations against genetic correlations across outcomes, to assess how well CRS effect correlation captured shared disease aetiology. By measuring the correlation in causal genetic effects between trait pairs, genetic correlation reflects the extent to which diseases share biological mechanisms at the molecular level ^27^. To compute the genetic correlations, we collected publicly available genome-wide association study (GWAS) summary statistics for 44 heritable diseases (out of the 88 outcomes) with heritability *z*-score > 6 that were primarily derived from European ancestry. These publicly available summary statistics were generated from (1) meta-analyses that included UK Biobank samples, or (2) Pan-UK Biobank study ^85^. We applied cross-trait linkage disequilibrium score regression (LDSC) ^86^ to the GWAS summary statistics for 1.2 million SNPs (minor allele frequency > 0.01) and used reference LD information from the 1000 Genomes ^87^ Europeans individuals (9,254,535 SNPs) to estimate genetic correlations. Details of phenotype information, heritability, and genetic correlation estimates from cross-trait LDSC for these 44 diseases are reported in Tables S31 and S6.

### Comparison with competing models

We compared CRS against state-of-the-art artificial intelligence (AI) models (Delphi-2M; tested outside the NHS SDE) ^1^, Phecode-based risk scores (PheRS) ^3^, state-of-the-art COVID-19 model (QCovid) ^4^, Charlson Comorbidity Index (CCI) ^20^, and socioeconomic status captured by Index of Multiple Deprivation (IMD) ^34^.

Delphi-2M is a GPT-based EHR model that simultaneously predicts over 1,000 diseases, trained using the UK Biobank (UKB) EHR data ^1^. Since the Delphi-2M model parameters were not publicly available (as compliant with UKB AI policies), we performed an indirect comparison of CRS evaluated in the CVD-COVID-UK cohort vs. Delphi-2M evaluated in the UKB cohort, by (1) matching the testing samples for CRS to have the same age range and observation timeframe as Delphi-2M under the longitudinal setting, and (2) matching Phecode-based CRS outcomes to ICD-10-based Delphi-2M outcomes based on Phecode Map 1.2 ^75^ (Table S13). Consistent with the Delphi-2M testing population, the testing samples for CRS in this comparison aged 50-80 at index and were followed up from 1 July 2021 to 1 July 2022, with predictor diagnoses evaluated up until 30 June 2020. The improvement in AUROC over the age-and-sex baseline model was compared between CRS and Delphi-2M for 49 overlapping outcomes, based on the testing performance of CRS in our cohort and longitudinal testing performance of Delphi-2M from Shmatko et al. ^1^ (Supplementary Fig. 4 of Shmatko et al., with source data provided by the authors).

PheRS is a Phecode-based elastic net model for predicting disease risk, trained using EHR data from multiple biobank studies including UKB ^3^. To compare CRS against PheRS, we implemented PheRS in the CVD-COVID-UK cohort using the PheRS effect sizes trained from the UKB data (Supplementary Table 12 of Detrois et al. ^3^). Phecode-based PheRS predictor diseases were identified from the combined data of HES APC and primary care. Out of the 13 outcomes considered in Detrois et al., 11 were selected and matched to CRS outcomes (Table S14), while major depression and epilepsy were excluded due to low incidence in our study population. For each outcome, the PheRS score was calculated in the training sample and calibrated in the same sample with age and sex as covariates, i.e., we fitted a model of Outcome ∼ PheRS + Age + Sex. The resulting calibrated PheRS model was then evaluated in the testing sample and compared directly against CRS in performance.

QCovid is a Cox proportional hazard model for predicting COVID-19 related mortality or hospitalisation, trained using the QResearch database ^10^. To compare CRS against QCovid for predicting COVID-19 hospitalisation, we implemented QCovid2, a version applicable to unvaccinated individuals (as our index date was before availability of COVID-19 vaccines), in the CVD-COVID-UK cohort. Consistent with previous implementation of QCovid in this cohort ^88^, we captured most predictor variables from QCovid, while excluding chemotherapy usage, housing status, and Townsend Deprivation Index as covariates due to data missingness. The full list of QCovid predictors along with effect sizes is provided in Table S32. QCovid was calculated in the training sample and retrained in two models: (1) COVID-19 hospitalisation ∼ QCovid (note that QCovid itself included age and sex as part of the model), and (2) COVID-19 hospitalisation ∼ CRS + QCovid + Age + Sex. These models were then evaluated and compared against CRS in the testing sample.

We further included CCI and IMD, respectively, as model covariates to assess the additional performance improvement by CRS over existing clinical comorbidity and socioeconomic indices. CCI was implemented based on Quan et al. ^20^, containing 12 comorbidities and extensively validated in previous administrative data. IMD was coded as quintiles and mapped to cohort individuals based on their lower layer super output areas (LSOAs) ^89^. For each outcome, we trained the following models in the training sample: (1) Outcome ∼ CCI + Age + Sex, (2) Outcome ∼ CRS + CCI + Age + Sex, (3) Outcome ∼ IMD + Age + Sex, and (4) Outcome ∼ CRS + IMD + Age + Sex. Model comparison was performed between (1) and (2), and between (3) and (4) in the testing sample.

### Cross-ethnicity and stratified evaluation

While CRSs were trained in White British individuals, we evaluated their performance in different self-reported ethnicity groups. Ethnicity groups were defined based on the 2011 UK Census ^90^. We tested CRSs’ performance in larger ethnicity groups based on the 5-group system: (1) Asian or Asian British (Asian), (2) Black, African, Caribbean or Black British (Black), (3) Mixed or multiple ethnic groups (Mixed), and (4) Irish or any other White background (Irish/Other White). Their sample sizes are reported in Fig. S1b-c.

To assess predictive performance of CRSs across longitudinal and demographic strata, trained CRS models were tested in subgroups of testing population stratified based on the following factors: (1) gap between predictor assessment and outcome incidence: 3 months (default), 1 year 3 months, 2 years 3 months, 4 years 3 months, 6 years 3 months, and 8 years 3 months; (2) age groups at index: 40-49, 50-59, and 60-69 years; and (3) sex: females and males.

### Comorbidity architecture of CRSs

We characterised the comorbidity architecture underlying the constructed CRSs by analysing the CRS effect sizes. Throughout this section, we assume that CRS effect sizes for each outcome *i*, 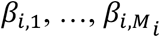 (Eq. (1)), have been standardised by predictor disease prevalence, i.e., the effect sizes are interpreted as log odds ratios associated with the outcome per standard deviation of the predictor. This allows us to compare the effect sizes of predictor diseases with different prevalences on the same scale. We also assume that 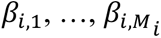 are independently sampled from a Gaussian distribution with mean 0.

First, we defined comorbidity architecture of a disease outcome as the distribution of CRS effect sizes on that outcome. Adapting a polygenicity measure from statistical genetics literature ^47^, we evaluated comorbidity architecture for each outcome *i* using the effective number of independently predictive diseases: *M*_*e*_ = 3*M*_*i*_/*κ*, where 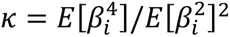. Intuitively, the *M*_*e*_ statistic captures how evenly the effect sizes are spread across predictor diseases: outcomes with many predictors contributing similar effect sizes will have a larger *M*_*e*_, indicating a higher level of multi-comorbidity; outcomes dominated by a few predictors with large effect sizes will have a smaller *M*_*e*_, indicating a lower level of multi-comorbidity. A naïve sample estimator of *M*_*e*_ can be constructed using sample moments of estimated effect sizes:

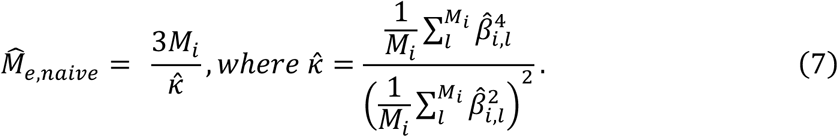

As sample moments are not unbiased estimators of population moments, we constructed the final estimator, *M̂*_*e*_, a modified version of *M̂*_*e*,*naive*_, by (1) using unbiased estimate of *κ*, and (2) using 2^nd^ order Taylor expansion to reduce bias caused by inverting *κ̂*.

We performed a simulation study to evaluate the accuracy of *M̂*_*e*_ in estimating *M*_*e*_ for a single outcome. To ensure realistic predictor prevalences and correlation structure, we generated a correlation matrix for predictor disease liability from observed pairwise correlations across the *M* = 212 predictor diseases in our training cohort. The resulting liability-scale correlation matrix along with observed predictor prevalences was used to generate simulated predictor data under a liability threshold model. Predictor effect sizes were simulated based on the following mixture distribution: for predictor *k =* 1,…, *M*, *β*_*k*_∼*Normal*(0, 0.03^2^) with a probability of *M*_*sim*_/*M*; *β*_*k*_ = 0 with a probability of 1 − *M*_*sim*_/*M*, with *M*_*sim*_ = 25, 50, 75, 100. Under such assumptions, the true *M*_*e*_ was equal to *M*_*sim*_ in each setting. Outcome data were generated based on logistic model-predicted risk from predictor data, with an average case number of 146,063 across 500 simulations for each *M*_*e*_. The *M̂*_*e*_ estimates from each simulation were then compared against the true *M*_*e*_ values.

Second, we quantified the relative contribution of each predictor disease and disease category (defined based on Phecode category) to the outcomes using their contribution to the *R*^2^ of the CRSs. The relative *R*^2^ contribution of prediction disease *k* to outcome *i* was defined as 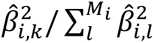 The relative *R*^2^ contribution of predictor category *C* (containing multiple predictor diseases) to outcome *i* was defined as 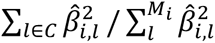

### Risk stratification and calibration of CRSs

For practical usage in clinical settings, we evaluated the performance of CRS in risk stratification. For each outcome, the entire testing population was divided into three groups based on their CRS score distribution: top 5%, middle 90%, and bottom 5%. In each group, Kaplan-Meier analysis was performed to estimate the cumulative disease risk over the study period, and the number of incident disease cases was recorded to evaluate CRSs’ ability in identifying individuals at high disease risk.

We further assessed the calibration of CRSs. As the training population in our main analysis had a 1:4 case-control ratio, not reflective of realistic disease incidence, we retrained the CRS models in a random subset of the training data with the original outcome incidence rate, with 3,000 cases, to obtain the correct model intercept corresponding to the real-world England population. The resulting updated model was then used to calculate CRS-predicted disease risk in the entire testing population. For each outcome, the testing population was divided into 20 equal quantiles based on the predicted risk from the model including CRS, age, and sex. We then compared the observed event rate within each quantile against the average predicted event rate within each quantile to evaluate the calibration performance of CRSs.

### Distinct predictive roles of comorbidities across outcomes revealed by CRS effect sizes

Lastly, we compared CRS effect sizes across outcomes to identify outcome pairs with high CRS effect correlation, and subsequently, predictor diseases with distinct roles in those outcomes. Standardised CRS effect sizes were used in this analysis. We focused on two examples in which CRS distinguished independently predictive comorbidities from indirect associations: (1) effect of tobacco use disorder (Phecode 318) on COVID-19 hospitalisation and sepsis (Phecode 994.2), and (2) effect of lipid metabolism disorder (Phecode 272) on myocardial infarction (Phecode 411.2) and ischaemic stroke (Phecode 433.21). To elucidate the mediative effects of other comorbidities on the predictor effect of interest (e.g., tobacco use disorder on COVID-19 hospitalisation), we estimated the effect size of the target predictor on the outcome while conditioning on different numbers of other comorbidities. To identify the top comorbidities that contribute to effect attenuation of the target predictor, we included one covariate comorbidity in the model at a time and ranked the resulting target predictor effect sizes based on magnitude of attenuation.

### Ethics approval

The North East - Newcastle and North Tyneside 2 research ethics committee provided ethical approval for the CVD-COVID-UK/COVID-IMPACT research programme (REC No 20/NE/0161) to access, within secure trusted research environments, unconsented, whole-population, anonymised data from EHRs collected as part of patients’ routine healthcare.

## Supporting information

SuppFigures

SuppTables

## Data Availability

The CVD-COVID-UK/COVID-IMPACT research programme, led by the BHF Data Science Centre (https://bhfdatasciencecentre.org) received approval to access data in NHS England's SDE service for England from the Advisory Group for Data (AGD) (https://digital.nhs.uk/about-nhs-digital/corporate-information-and-documents/advisory-group-for-data) -- formerly the Independent Group Advising on the Release of Data (IGARD) -- via an application made in the Data Access Request Service (DARS) Online system (ref. DARS-NIC-381078-Y9C5K) (https://digital.nhs.uk/services/data-access-request-service-dars/dars-products-and-services). The CVD-COVID-UK/COVID-IMPACT Approvals & Oversight Board (https://bhfdatasciencecentre.org/areas/cvd-covid-uk-covid-impact/) subsequently granted approval to this project (CCU022) to access the data within NHS England's SDE service for England. The anonymised data used in this study were made available to accredited researchers only. Those wishing to gain access to the data should follow the application process of the relevant national data custodian.

## Acknowledgments

This work was carried out with the support of the BHF Data Science Centre led by Health Data Research UK (BHF Grant no. SP/19/3/34678). This study made use of anonymised data held in NHS England’s Secure Data Environment service for England and made available via the BHF Data Science Centre’s CVD-COVID-UK/COVIDIMPACT consortium. This work used data provided by patients and collected by the NHS as part of their care and support. We would also like to acknowledge all data providers who make health relevant data available for research.

The BHF Data Science Centre’s Health Data Science Team provided data curation resources and support. We would also like to acknowledge Dr. Genevieve Cezard for providing guidance on QCovid score computation.

## Funding

Wellcome Trust early-career award no. 227566/Z/23/Z (XJ)

National Institutes of Health grants R01 HG006399 (ALP) and R37 MH107649 (ALP)

The British Heart Foundation Data Science Centre (grant No SP/19/3/34678, awarded to Health Data Research (HDR) UK) funded co-development (with NHS England) of the Secure Data Environment service for England, provision of linked datasets, data access, user software licences, computational usage, and data management and wrangling support, to coordinate national COVID-19 priority research. Consortium partner organisations funded data access, user software licences and computational usage, and the time of contributing data analysts, biostatisticians, epidemiologists, and clinicians.

## Author contributions

Conceptualization: ALP, XJ, HL

Resources: XJ, AW, MI

Data curation: MAM, HL

Methodology: HL, YZ, XJ

Formal analysis: HL, YZ, XJ

Software: HL

Funding acquisition: XJ

Supervision: XJ

Writing – original draft: HL, XJ, ALP

Writing – review & editing: HL, MAM, YZ, AW, MI, ALP, XJ

## Competing interests

Authors declare that they have no competing interests.

## Data, code, and materials availability

We have developed the “crscalc” R package to facilitate practical implementation of CRSs, with instructions for usage available from: https://github.com/pearl-liu/crscalc.

The data used in this study are available in NHS England’s SDE service for England, but as restrictions apply, they are not publicly available (https://digital.nhs.uk/services/secure-data-environment-service). The CVD-COVID-UK/COVID-IMPACT research programme, led by the BHF Data Science Centre (https://bhfdatasciencecentre.org) received approval to access data in NHS England’s SDE service for England from the Advisory Group for Data (AGD) (https://digital.nhs.uk/about-nhs-digital/corporate-information-and-documents/advisory-group-for-data) – formerly the Independent Group Advising on the Release of Data (IGARD) – via an application made in the Data Access Request Service (DARS) Online system (ref. DARS-NIC-381078-Y9C5K) (https://digital.nhs.uk/services/data-access-request-service-dars/dars-products-and-services). The CVD-COVID-UK/COVID-IMPACT Approvals & Oversight Board (https://bhfdatasciencecentre.org/areas/cvd-covid-uk-covid-impact/) subsequently granted approval to this project (CCU022) to access the data within NHS England’s SDE service for England. The anonymised data used in this study were made available to accredited researchers only. Those wishing to gain access to the data should follow the application process of the relevant national data custodian.

The study protocol, analysis code, phenotype definitions, and code lists will be made available by the BHF Data Science Centre through GitHub repository before publication (currently under review).

## Supplementary materials

Figs. S1 to S12

Tables S1 to S32 (combined into a single Excel file as Data S1)

Data S1

## Notes

### Competing Interest Statement

The authors have declared no competing interest.

### Summary of Updates

Resubmitting PDF file instead of word file as the previous figure resolution was really low during the conversion.

