## Supplementary material for "Predicting COVID-19 hospitalisation and common disease risk from comorbid diagnoses in 13 million individuals": SuppFigures

**The PDF file includes:**

Materials and methods

Figs. S1 to S12

Tables S1 to S32 (captions only)

**Other Supplementary Materials for this manuscript include the following:**

Data S1

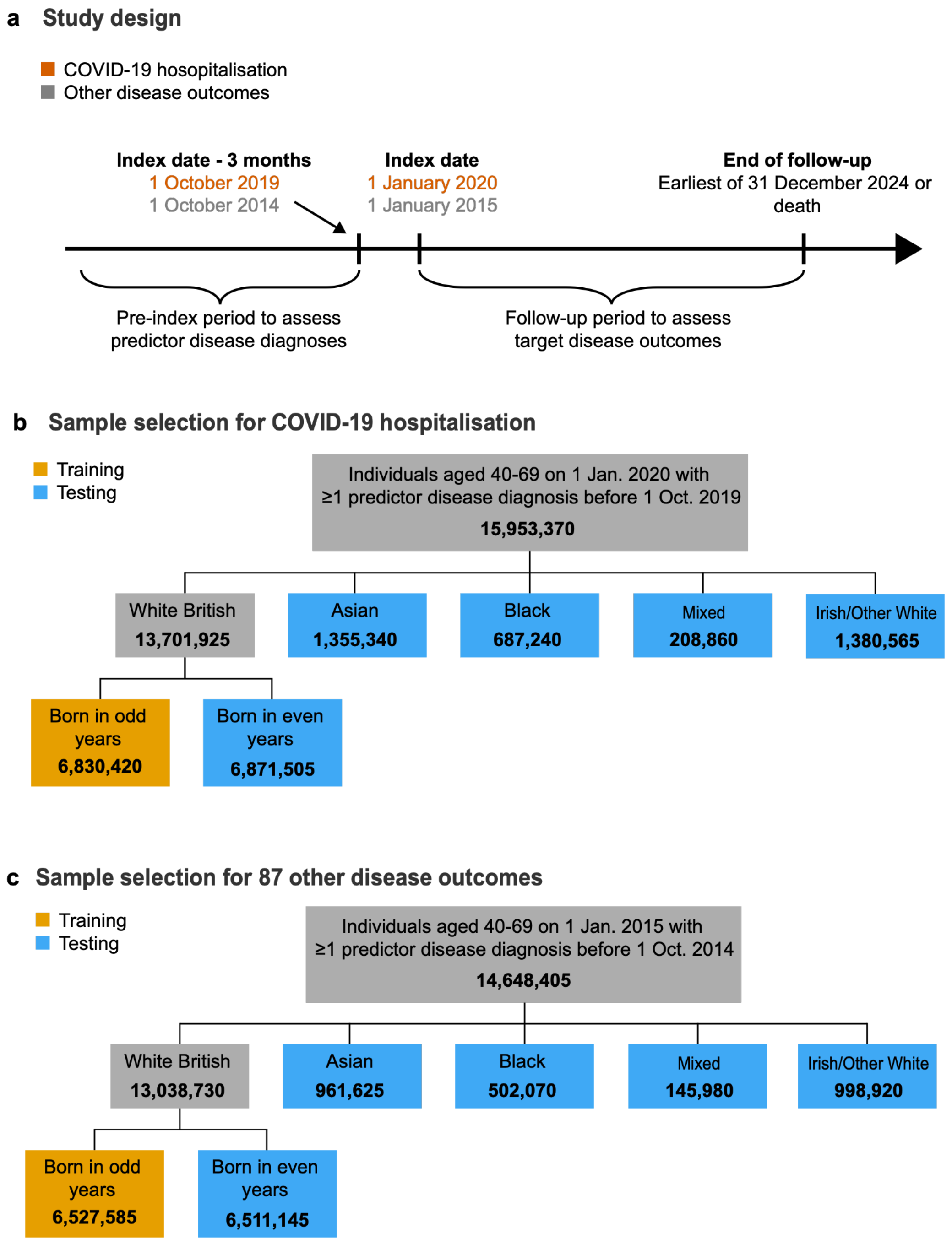


**Fig. S1.**

**Overview of study design and sample selection. a.** Schematic overview of study design for predicting COVID-19 hospitalisation and 87 other common disease outcomes in CVD-COVID-UK. **b.** Sample selection of training and testing cohorts for COVID-19 hospitalisation. Sample sizes (rounded to the nearest multiple of 5) are reported at each step. **c.** Sample selection of training and testing cohorts for 87 other disease outcomes. Sample sizes (rounded to the nearest multiple of 5) are reported at each step.

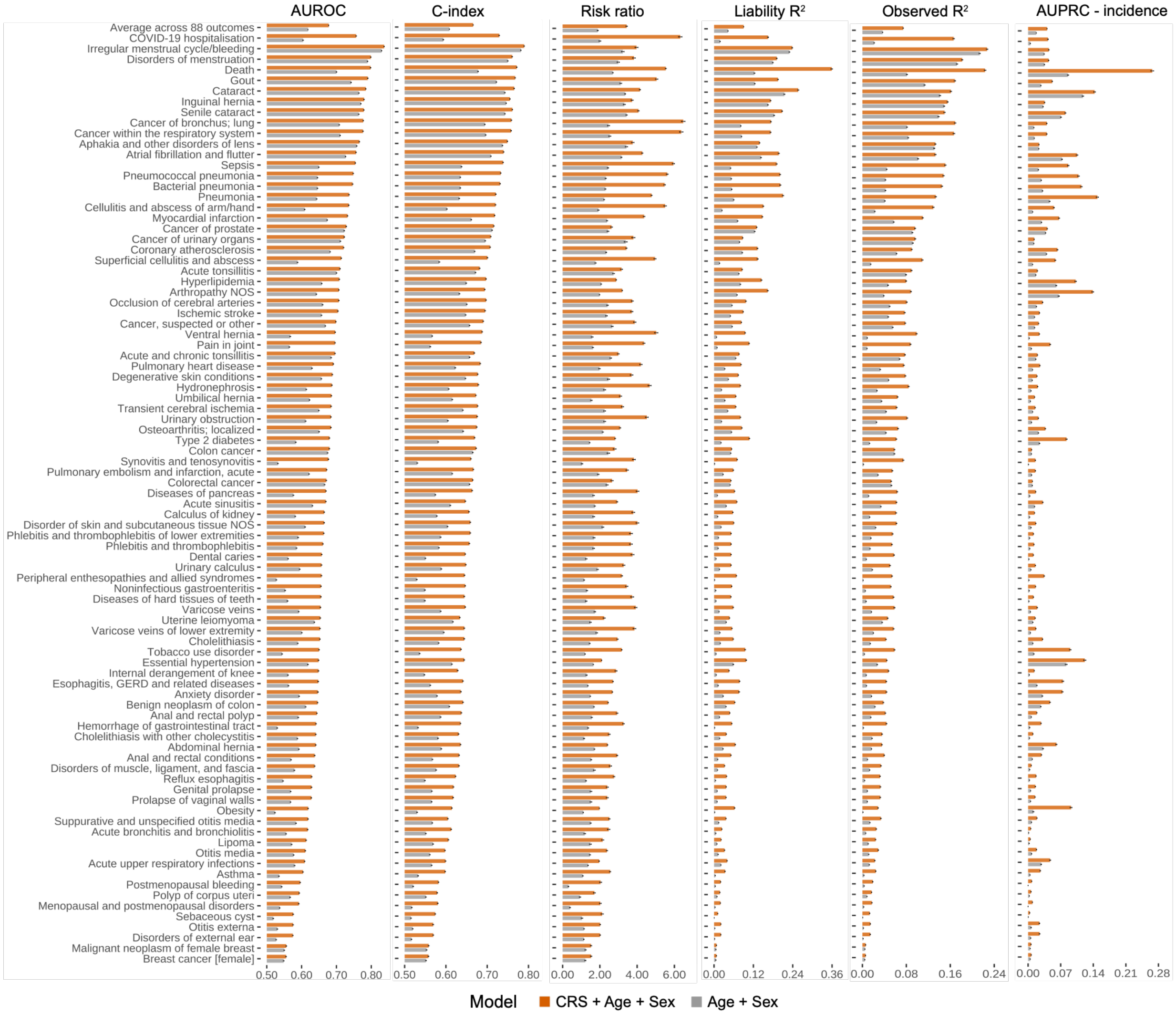


**Fig. S2.**

**CRS predictive performance for 88 outcomes evaluated using six metrics.** AUROC, C-index, observed risk ratio (top 5% of the predicted risk distribution vs. population average), liability-scale $R^2$, observed-scale $R^2$, and the difference between AUPRC and outcome incidence for all 88 outcomes are shown for the CRS model (including age and sex as covariates) vs. a baseline age-and-sex model. Error bars represent 95% CI around the performance metrics (but are generally very narrow). Following average performance and COVID-19 hospitalisation, the outcomes are ordered based on their AUROC values by the CRS model.

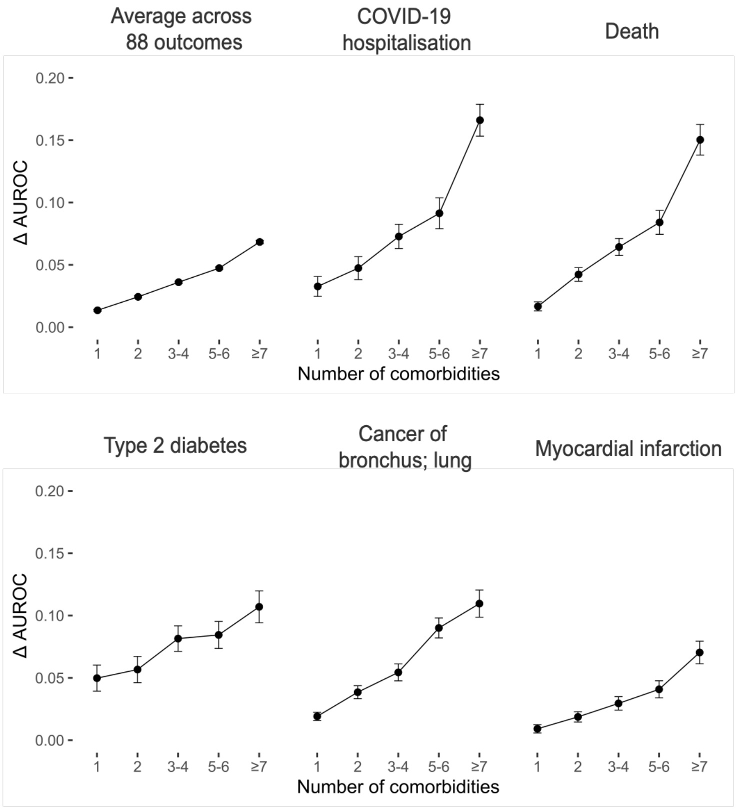


**Fig. S3.**

**CRS predictive performance stratified by number of comorbidities.** Improvement in AUROC by CRS over the age-and-sex baseline (ΔAUROC; *y*-axis) vs. individuals' number of comorbidities (*x*-axis) is shown for example outcomes. Performance was evaluated within each stratum that contained individuals with the target number of comorbidities. Error bars represent 95% CI around ΔAUROC (very narrow for the average across 88 outcomes).

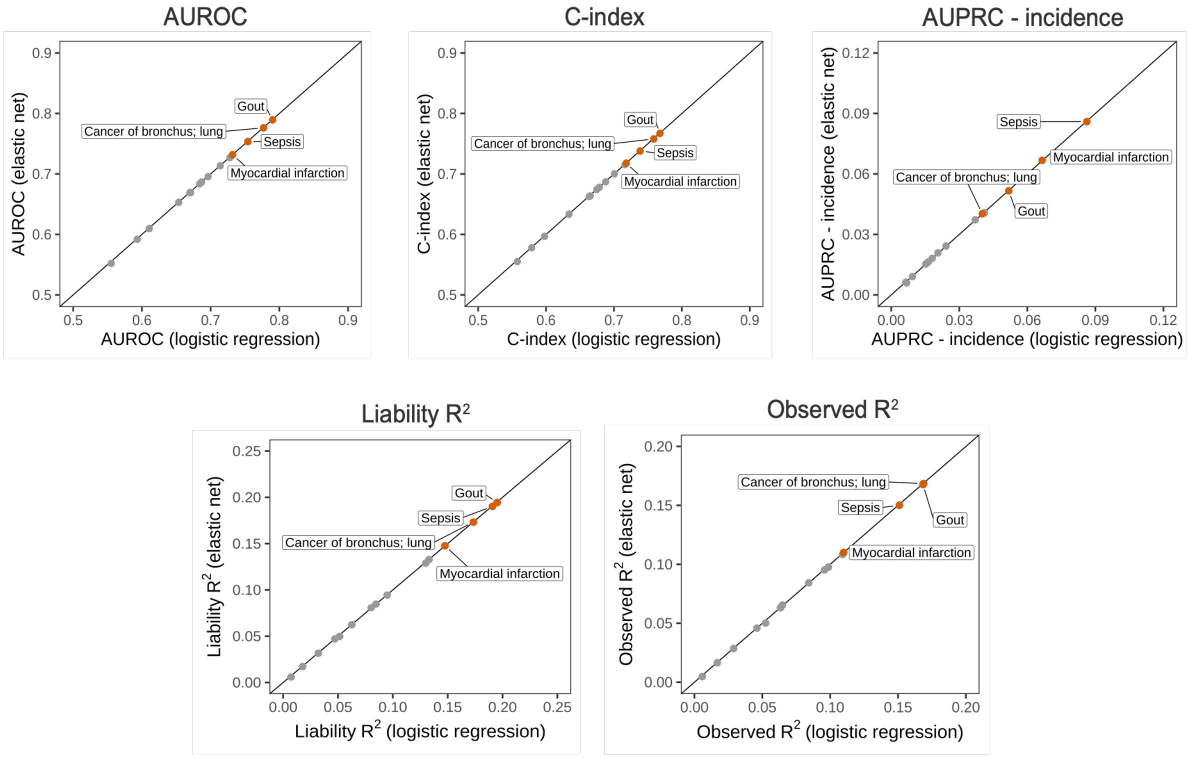


**Fig. S4.**

**Predictive performance of elastic net-based CRS vs. logistic regression-based CRS.** AUROC, C-index, the difference between AUPRC and outcome incidence, liability-scale $R^2$, and observed-scale $R^2$ are compared between elastic net-based CRS model ($y$-axis) and logistic regression-based CRS model (default model; $x$-axis) for 15 outcomes. In elastic net model training, the mixing parameter (alpha) and the regularisation strength (lambda) were simultaneously optimised using grid search and 10-fold cross-validation (via cva.glmnet function from glmnetUtils R package). The following outcomes were included in this comparison to capture a range of performances while limiting the computational burden of elastic net model fitting: Phecodes 153, 165.1, 174.1, 185, 218.1, 274.1, 381.1, 411.2, 550.5, 577, 595, 622.1, 681, 740.1, and 994.2.

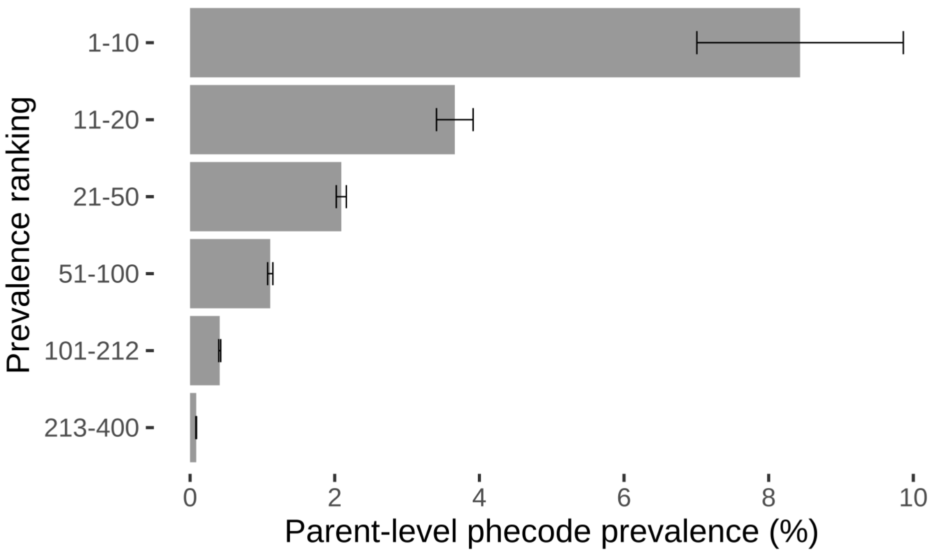


**Fig. S5.**

**Prevalence of parent-level Phecodes in CVD-COVID-UK.** The average prevalence of parent-level Phecodes (*x*-axis) is shown for Phecode groups (*y*-axis) defined based on their prevalence ranking (highest to lowest, i.e., top 1-10 prevalent Phecodes, top 11-20 prevalent Phecodes, etc.). Prevalences were calculated as of 1 January 2015, among individuals aged 40-69 on 1 January 2015 in the combined data of primary care + HES APC. Error bars represent standard errors around the mean prevalence, averaged across Phecodes in each group.

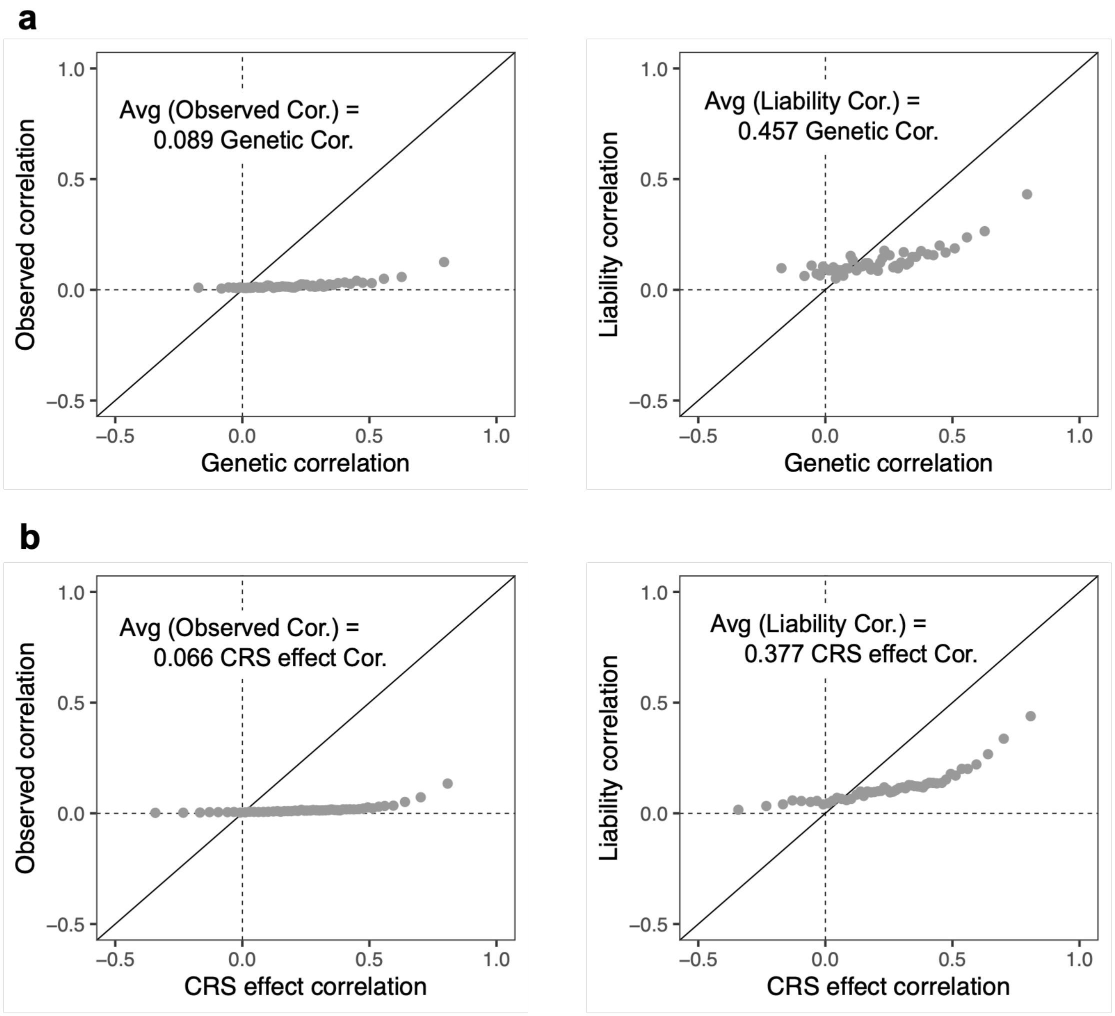


**Fig. S6.**

**Comparison of different types of outcome correlations. a.** Observed-scale and liability-scale phenotypic correlations (*y*-axis) vs. genetic correlation (*x*-axis) across outcomes. **b.** Observed-scale and liability-scale phenotypic correlations (*y*-axis) vs. CRS effect correlation (*x*-axis) across outcomes. In each panel, the pairwise correlation on the *x*-axis was divided into 50 equal quantiles, and the mean observed/liability correlations within each quantile are plotted against the mean genetic or CRS effect correlation within each quantile. The slope from a linear regression through the origin across all pairs of outcomes ($N = 936$ for Panel **a**; $N = 3{,}802$ for Panel **b**) is reported in text.

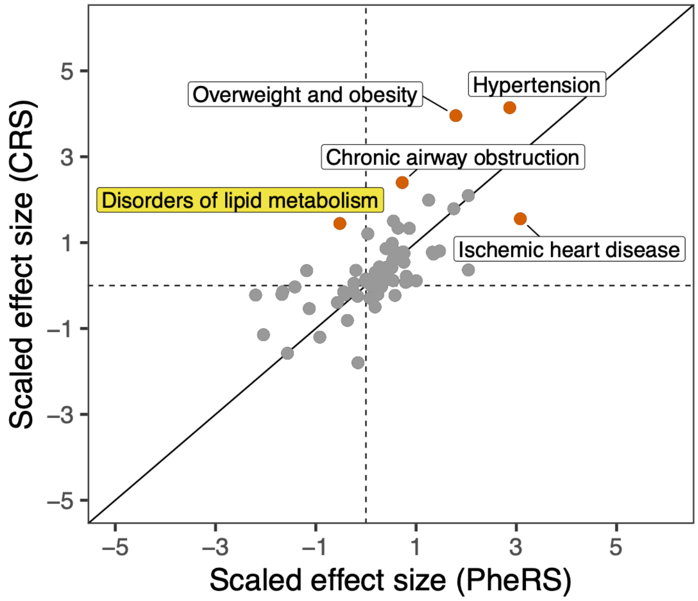


**Fig. S7.**

**Predictor effect size comparison between CRS and PheRS for Type 2 diabetes.** As CRS and PheRS effect sizes have different variances, for better visualisation, scaled CRS effect sizes (log odds ratio divided by its standard deviation; *y*-axis) is plotted against scaled PheRS effect sizes (*x*-axis) for overlapping predictors. Five predictors with large CRS effect sizes are labelled and highlighted in red points. Disorders of lipid metabolism, with positive CRS effect size but weak negative PheRS effect size, is further highlighted in yellow label.

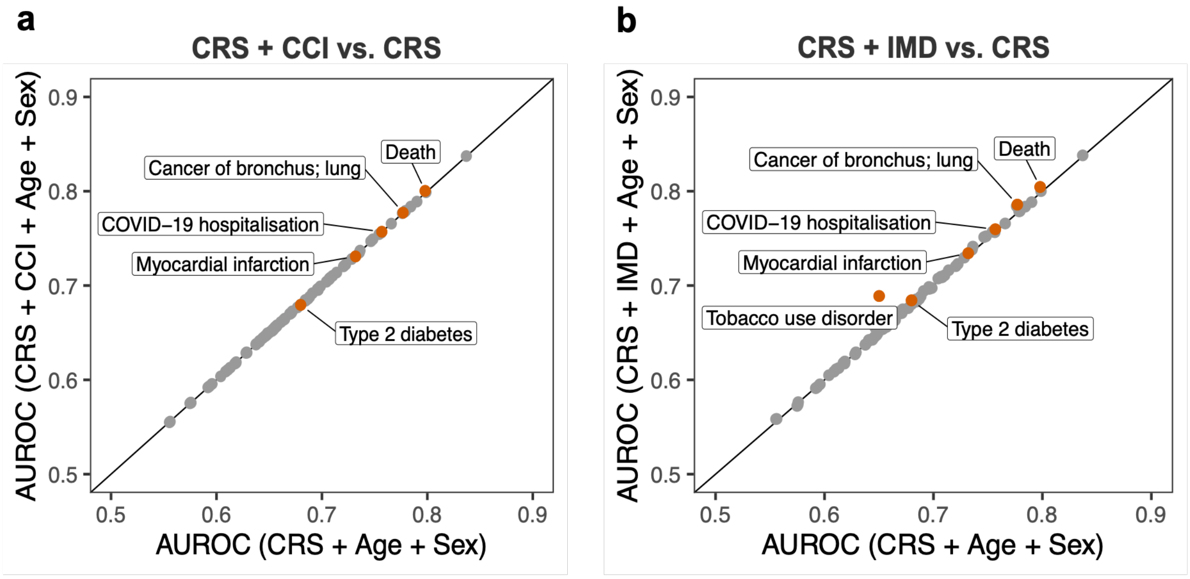


**Fig. S8.**

**Improvement in CRS by including CCI or IMD as a covariate. a.** AUROC for the model including CRS, CCI, age, and sex (*y*-axis) vs. AUROC for the model including CRS, age, and sex (*x*-axis). **b.** AUROC for the model including CRS, IMD, age, and sex (*y*-axis) vs. AUROC for the model including CRS, age, and sex (*x*-axis). All 88 outcomes are shown, with example outcomes labelled and highlighted in red points.

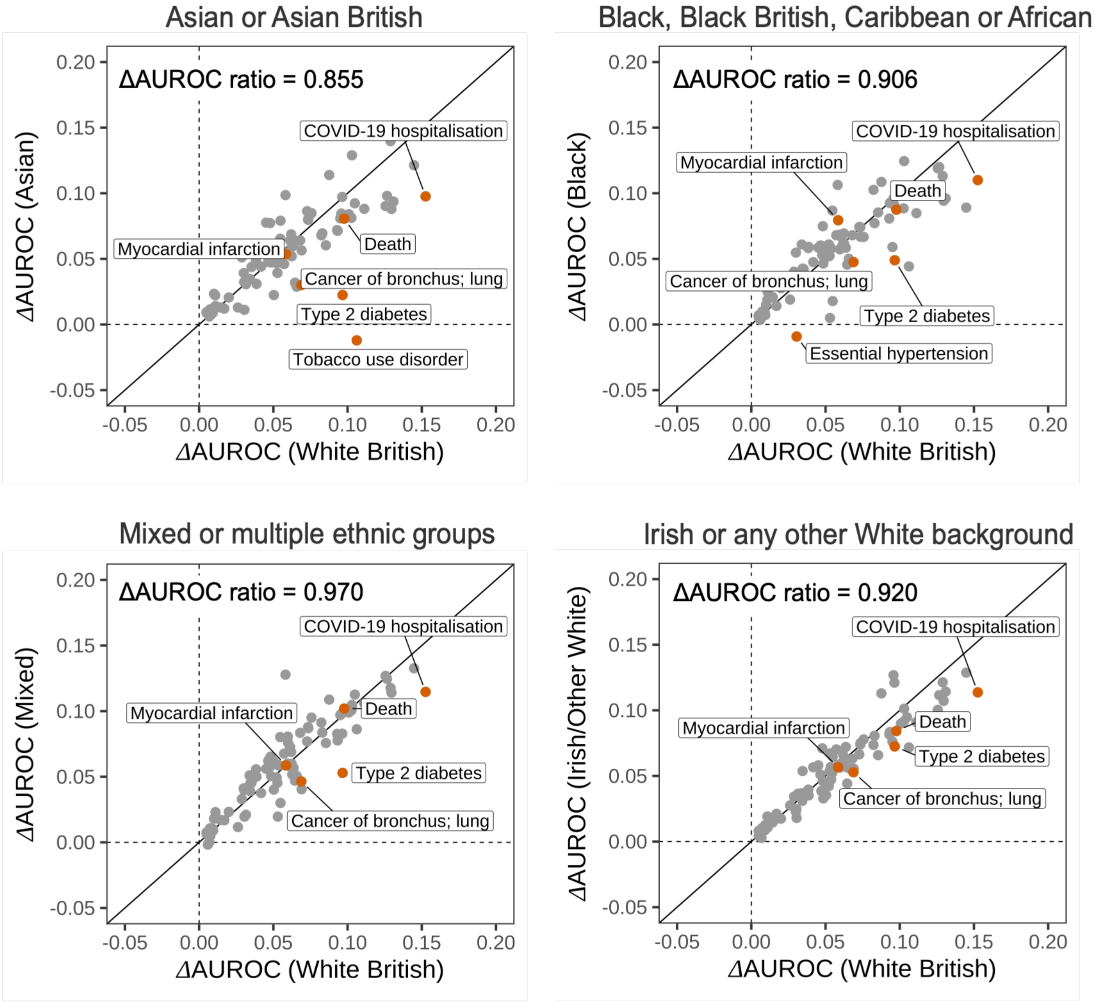


**Fig. S9.**

**Cross-ethnicity testing performance of CRS trained in White British individuals, assessed by ΔAUROC.** **Δ**AUROC (CRS vs. age-and-sex baseline) assessed in held-out Asian, Black, Mixed, and Irish/Other White ethnicity groups (*y*-axis) are separately compared against **Δ**AUROC assessed in held-out White British group (*x*-axis). All 88 outcomes are shown, with example outcomes labelled and highlighted in red points. In each comparison, the slope of a linear regression through the origin is reported in text.

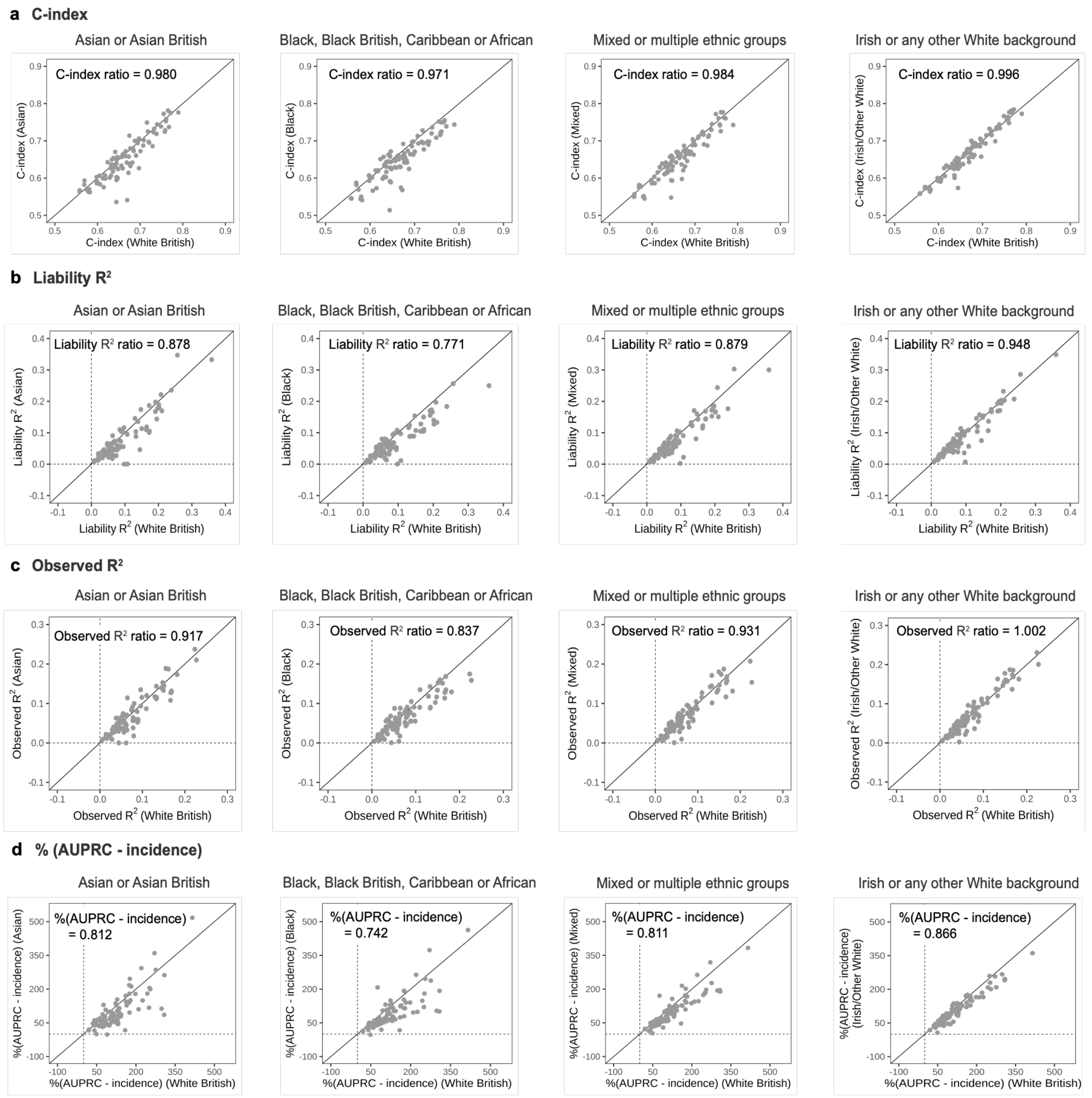


**Fig. S10.**

**Cross-ethnicity testing performance of CRS trained in White British individuals, assessed by four different metrics**. C-index (**a**), liability-scale $R^2$ (**b**), observed-scale $R^2$ (**c**), and percentage of difference between AUPRC and outcome incidence (**d**) assessed in held-out Asian, Black, Mixed, and Irish/Other White ethnicity groups (*y*-axis) are separately compared against each metric assessed in held-out White British group (*x*-axis). Percentage of AUPRC-incidence difference was calculated as (AUPRC - incidence)/incidence*100; this metric was used instead of the raw difference to account for different disease incidences of the same outcome across different ethnicity groups. All 88 outcomes are shown. In each comparison, the slope of a linear regression through the origin is reported in text.

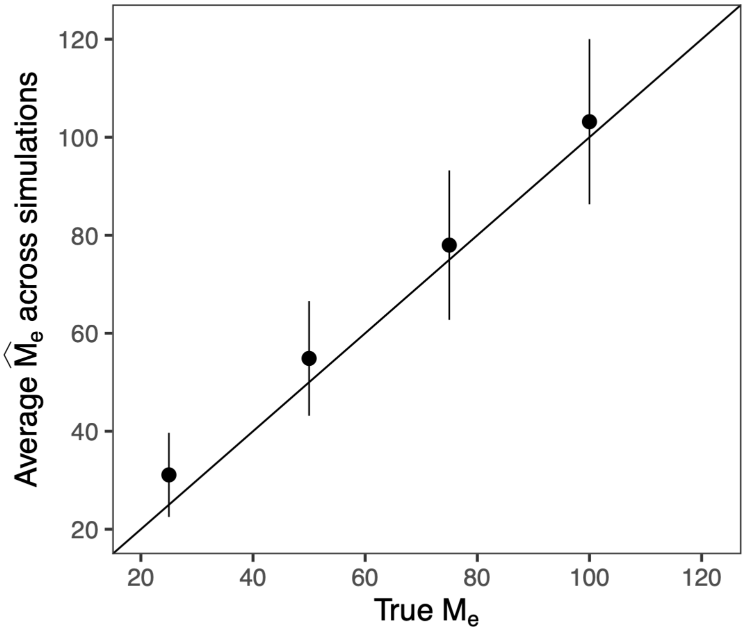


**Fig. S11.**

**Simulation performance of $\widehat{M}_e$ as an estimator of the effective number of independently predictive comorbidities**. The average $\widehat{M}_e$ estimate across 500 simulations ($y$-axis) is compared against the true $M_e$ value ($x$-axis) in each simulation setting. Error bars on the $y$-axis represent empirical standard errors around $\widehat{M}_e$ estimates across simulations. We note that, in practice, we estimated the standard errors using delta method, which was slightly anti-conservative with an empirical 95% CI coverage of 90% on average.

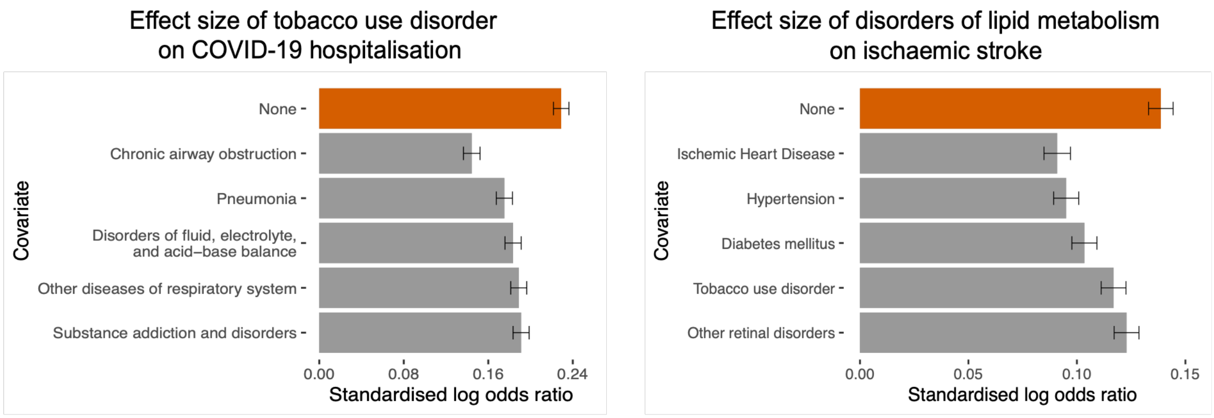


**Fig. S12.**

**Effect size attenuation of (1) tobacco use disorder on COVID-19 hospitalisation, and (2) disorders of lipid metabolism on ischaemic stroke.** For each target association**,** covariate comorbidities were included in the regression model one at a time. The resulting effect sizes (standardised log odds ratio) of the target predictor are shown (grey), along with the marginal effect size (red) when no covariate disease is included. The top 5 covariate diseases that caused the largest effect size attenuation are reported in each panel. Error bars represent 95% CI around the effect sizes.

Tables S1-S32 are combined into a single Excel file as Data S1. Here only table captions are described.

**Table S1.**

**Description of 88 disease outcomes.** The following outcome information is reported: Phecodes, phenotype definitions, Phecode categories, applicable sex, number of training cases, incidence in the training data, and sex-specific incidence in the training data (for sex-specific outcomes). Cases and incidences were calculated during the 5-year follow-up period (2020-2024) for COVID-19 hospitalisation and during the 10-year follow-up period (2015-2024) for other outcomes.

**Table S2.**

**Description of 212 predictor diseases.** The following predictor information is reported: Phecodes, phenotype definitions, Phecode categories, and prevalences in the training data as of index dates, 1 January 2015 and 1 January 2020, respectively.

**Table S3.**

**CRS effect size estimates trained in the full cohort of White British individuals.** The summary statistics were derived from IPCW-weighted logistic regression with robust standard errors, with a control-case ratio of up to 4:1 for each outcome. Key columns include: outcome Phecode and definition, predictor Phecode and definition, CRS raw effect size estimates (i.e., log odds ratio; "Coef_Est") and standard errors ("Coef_SE"), CRS effect size estimates standardised by predictor disease prevalences ("Coef_Est_PredictorStandardised"), CRS effect size p-values ("P_value"), and false discovery rate-adjusted p-values ("P_value_FDR").

**Table S4.**

**CRS predictive performance for 88 outcomes.** Six metrics are reported: AUROC ("AUROC"), C-index ("C_index"), observed risk ratio ("RR_Top5perc_vs_Avg"; top 5% of the predicted risk distribution vs. population average), liability-scale $R^2$ ("R2_liab"), observed-scale $R^2$ ("R2_obs"), and the difference between AUPRC and outcome incidence ("AUPRC_incid_diff"). Key columns include: outcome Phecode and definition, model type ("Model"; CRS + Age + Sex vs. Age + Sex), point estimate of each metric (with "_mean" suffix), 95% CI of each metric (with "_LB" and "_UB" suffixes for lower and upper bounds), difference in metric between CRS and age-and-sex baseline (with "Delta_" prefix), and p-value for the difference between CRS and age-and-sex baseline (with "Delta_" prefix and "_Pval" suffix). Note that "Delta_RR_Top5perc_vs_Avg" represents the observed risk ratio of top 5% of CRS-predicted risks vs. top 5% of age-and-sex-predicted risks. Source data for Fig. 1a.

**Table S5.**

**CRS effect size estimates trained in the training cohort of White British individuals.** The training cohort included White British individuals born in odd years. The summary statistics were derived from IPCW-weighted logistic regression with robust standard errors, with a control-case ratio of up to 4:1 for each outcome. Key columns include: outcome Phecode and definition, predictor Phecode and definition, CRS raw effect size estimates (i.e., log odds ratio; "Coef_Est") and standard errors ("Coef_SE"), CRS effect size estimates standardised by predictor disease prevalences ("Coef_Est_PredictorStandardised"), CRS effect size p-values ("P_value"), and false discovery rate-adjusted p-values ("P_value_FDR").

**Table S6.**

**CRS effect correlation, phenotypic correlation, and genetic correlation across outcomes.** Different types of correlation are reported for each eligible pair of outcomes, with the following key columns: outcome 1 ("Outcome_Phecode1"), outcome 2 ("Outcome_Phecode2"), CRS effect correlation ("CRS_effect_corr") and standard error ("CRS_effect_corr_SE"), observed-scale phenotypic correlation ("Observed_corr"), liability-scale phenotypic correlation ("Liability_corr"), whether genetic correlation is available for the outcome pair ("Genetic_corr_available"), source GWAS summary statistics for calculating genetic correlation ("GeneticSource1__GeneticSource2"), genetic correlation ("Genetic_corr"), and its standard error ("Genetic_corr_SE"). Source data for Fig. 1e.

**Table S7.**

**Stratified CRS predictive performance for 88 outcomes.** Five metrics are reported: AUROC ("AUROC"), C-index ("C_index"), liability-scale $R^2$ ("R2_liab"), observed-scale $R^2$ ("R2_obs"), and the difference between AUPRC and outcome incidence ("AUPRC_incid_diff"). Key columns include: outcome Phecode and definition, model type ("Model"; CRS + Age + Sex vs. Age + Sex), stratifying variable ("Stratified_by"), strata definition ("Strata_value"), point estimate of each metric (with "_mean" suffix), 95% CI of each metric (with "_LB" and "_UB" suffixes for lower and upper bounds), difference in metric between CRS and age-and-sex baseline (with "Delta_" prefix), and p-value for the difference between CRS and age-and-sex baseline (with "Delta_" prefix and "_Pval" suffix). Source data for Fig. 3b-d.

**Table S8.**

**Observed outcome risk among the top 5% risk group predicted by CRS vs. the top 5% risk group predicted by age and sex.** For each outcome, observed risks were estimated by the end of the 10- or 5-year follow-up period using Kaplan-Meier method. Key columns include: outcome Phecode and definition, observed risk in the top 5% risk group predicted by CRS ("Observed_risk_Top5perc_CRS") and its standard error ("Observed_risk_Top5perc_SE_CRS"), observed risk in the top 5% risk group predicted by age and sex ("Observed_risk_Top5perc_Age_Sex"), and its standard error ("Observed_risk_Top5perc_SE_Age_Sex").

**Table S9.**

**Calibrated CRS model coefficients trained in the training cohort of White British individuals.** The CRS models were calibrated in the training cohort with realistic outcome incidence, based on effect size estimates from **Table S5**. Key columns include: outcome Phecode and definition, predictor Phecode and definition, and CRS effect size estimates (i.e., log odds ratio; "Coef_Est"). Note that, compared to **Table S5**, the calibration only substantially changed the intercept in each outcome model, but the predictor effect estimates remained similar due to properties of log odds ratio.

**Table S10.**

**Liability $R^2$ in the testing data vs. liability $R^2$ in the training data.** Source data for Fig. 1b.

**Table S11.**

**Improvement in liability $R^2$ by CRS over the age-and-sex baseline vs. number of predictor diseases included in the CRS model.** Results for all 88 outcomes are reported in this table. Source data for Fig. 1c.

**Table S12.**

**Effect sizes (log odds ratio) of the full CRS model with 212 predictors vs. effect sizes of the CRS model with top 100 prevalent predictors.** Source data for Fig. 1d.

**Table S13.**

**Improvement in AUROC (ΔAUROC) by CRS over the age-and-sex baseline vs. ΔAUROC by Delphi-2M over the age-and-sex baseline.** Source data for Fig. 2a.

**Table S14.**

**Improvement in AUROC (ΔAUROC) by CRS over the age-and-sex baseline vs. ΔAUROC by PheRS over the age-and-sex baseline.** Source data for Fig. 2b.

**Table S15.**

**Improvement by CRS over existing clinical risk models in predicting COVID-19 hospitalisation.** Source data for Fig. 2c.

**Table S16.**

**Improvement by CRS over CCI for all 88 outcomes.** Source data for Fig. 2d.

**Table S17.**

**Improvement by CRS over IMD for all 88 outcomes.** Source data for Fig. 2e.

**Table S18.**

**Cross-ethnicity testing performance of CRS trained in White British individuals.** Source data for Fig. 3a.

**Table S19.**

**Effective number of independently predictive diseases, $\widehat{M}_e$, for all 88 outcomes.** Source data for Fig. 4a.

**Table S20.**

**Percentage of CRS $R^2$ contribution from predictor categories as defined by Phecode categories for all 88 outcomes.** Source data for Fig. 4b.

**Table S21.**

**Percentage of CRS $R^2$ contribution from each predictor disease averaged across 88 outcomes.** Source data for Fig. 4c.

**Table S22.**

**Percentage of CRS $R^2$ contribution from each predictor category averaged across 88 outcomes.** Source data for Fig. 4d.

**Table S23-S27.**

**Summary statistics from Kaplan-Meier analyses of cumulative event rate for COVID-19 hospitalisation (S23), death (S24), lung cancer (S25), myocardial infarction (S26), and type 2 diabetes (S27), stratified by CRS risk groups.** Source data for Fig. 5a.

**Table S28.**

**Calibration results of CRS for all 88 outcomes.** Source data for Fig. 5b.

**Table S29.**

**CRS effect sizes for (1) sepsis vs. COVID-19 hospitalisation, and (2) myocardial infarction vs. ischaemic stroke.** Source data for Fig. 6 (left panels).

**Table S30.**

**Effect sizes of target predictor vs. number of covariate predictor diseases included in CRS for target outcomes.** Source data for Fig. 6 (right panels).

**Table S31.**

**Phenotype and heritability information for 44 outcome diseases used to calculate genetic correlation.** Key columns include: outcome Phecodes and phenotypes, source GWAS summary statistics ("Genetic_Source"), heritability estimate ("Heritability_Est"), standard error ("Heritability_SE"), and z-score ("Heritability_Z"). Note that one outcome can have multiple summary statistics sources; in the genetic correlation analysis, the pair of summary statistics that produced the lowest standard error for genetic correlation was chosen for the target pair of outcomes.

**Table S32.**

**Predictor coefficients for QCovid2 model.** This table includes QCovid2 predictor definitions, predictor coefficients (log hazard ratios) for females, predictor coefficients for males, and indicator of whether the predictor was used in the analysis of CVD-COVID-UK data.

**Data S1. (separate file)**

An Excel file including Tables S1-S32, with each supplementary table presented in separate sheets. The sheet name corresponds to table numbers.
